# Anxiety associated with dietary intake and gut microbiome features in a cross-sectional cohort of sub-clinically anxious young women

**DOI:** 10.64898/2026.03.18.26348688

**Authors:** Melissa Basso, Falk Hildebrand, Chantelle Winder, David J Baker, Ralph Manders, Matteo Barberis, Sean M. Gibbons, Kathrin Cohen Kadosh

## Abstract

**Background:** Emerging evidence highlights the gut-brain axis as a key pathway linking diet and anxiety, yet the key determinants remain unclear. Most studies have focused on single components of diet and rarely integrate long- and short-term intake. Furthermore, prior gut-brain work has focused on microbiome composition, while functional features remain underexplored. In this study, we investigated associations between long- and short-term dietary intake, gut microbiome composition and functions, and anxiety in a subclinical cohort of 46 females (18–24 years) from the United Kingdom.

**Results:** Long-term diet quality was assessed using the Healthy Eating Index (HEI-2020) derived from a food frequency questionnaire, stratifying participants into lower and higher diet quality clusters. Short-term dietary intake was assessed via 24-hour recalls. Shotgun metagenomics of stool samples was used to assess differences in α and β diversity indices, species abundances, and bacterial pathways putatively metabolizing gut-brain-axis relevant molecules. Anxiety was measured using the State–Trait Anxiety Inventory (state subscale – STAI-s). Regression models identified diet quality (HEI cluster) as the primary dietary feature of anxiety variation. The presence of *Ruminococcus B gnavus* and *Flavonifractor plautii*, and the abundances of *Bilophila wadsworthia* and *Bacteroides thetaiotaomicron* were positively associated with anxiety. The presence of *Feacalibacterium prausnitzii* and greater abundances of butyrate, propionate, and GABA synthesis pathways were inversely associated with anxiety. Non-linear models revealed a U-shaped relationship between inositol synthesis and STAI-s. Finally, we found that habitual diet quality may modulate anxiety-related responses to short-term dietary variation.

**Conclusions:** These findings reveal widespread links between long-term diet quality, microbiota composition and function, and anxiety symptoms. These results point towards several promising targets for prebiotic, probiotic, postbiotic, and dietary interventions targets aimed at reducing anxiety.

## 1. Introduction

Anxiety has a lifetime global prevalence of 33%, with deleterious effects on quality of life and increased risk for cardiovascular and neurodegenerative diseases (1,2). For example, a recent study showed a 2.8-fold greater risk of Alzheimer disease over 10-year-follow-up in individuals with clinical anxiety (3). Panic and generalized anxiety disorders typically emerge in early adulthood and midlife, with higher prevalence in females (4,5). Despite treatment, these disorders can become chronic, with about 40% of patients remaining treatment-resistant (6). A meta-analysis reported only limited effect sizes of psychotherapies and pharmacotherapies, a high risk of study bias (7), and a 12-year prospective study found low recovery and high symptom recurrence rates (8).

Growing evidence links diet to anxiety risk, highlighting a promising, low-cost avenue for prevention and treatment. For example, consistent with Lee et al. (9), our recent review found that pro-inflammatory diets and diets rich in ultra-processed foods (UPFs) were most strongly associated with increased anxiety, whereas Mediterranean and other healthy dietary patterns were associated with reduced anxiety (10). However, most studies examine only one hierarchical level of diet (e.g., nutrients, foods, or quality indices) making it difficult to pinpoint the major dietary drivers of anxiety risk or symptom variability (10).

Diet acts as a substrate for gut microbes and may influence anxiety through gut microbiome-associated mechanisms. The gut-brain axis has been implicated in anxiety symptomatology, mediated systemically and/or peripherally via immune, endocrine, autonomic, and metabolic pathways (10). For example, a prebiotic intervention in mice with fructo- and galacto-oligosaccharides reduced anxiety-like behaviour and stress-induced corticosterone release, and increased serotonin levels in the prefrontal cortex, possibly through alteration of short-chain-fatty-acids (SCFAs) production in the gut (11). In humans, studies linking the gut microbiome to anxiety are still limited and report heterogeneous, sex- and population-specific effects (10,12,13). Similarly pre- and probiotic interventions showed inconsistent results, likely due to variation in study design, operationalization strategies, participant populations, dosage, strain, or substrate type, and, critically, uncontrolled individual-specific differences in diet and other lifestyle factors (13,14).

To address these gaps, we have proposed a new theoretical and methodological framework in which the gut microbiome may mediate the diet-anxiety relationship, emphasizing: i) hierarchical approaches to diet conceptualization and operationalization, ii) advancing mechanistic insights through the integration of community-level and functional/metabolic gut microbiome data; and iii) adopting a multifactorial, systems-level approach wherever feasible (10). This is in line with recent considerations for microbiome trials suggested by the International Scientific Associations for Probiotics and Prebiotics (ISAPP) (15).

Furthermore, it is important to distinguish between ‘trait’ and ‘state’ anxiety. Trait refers to a relatively stable predisposition shaped by genetic factors, life-long experiences, and behavioural patterns. State indicates a transient condition, in response to the internal and external environment (16,17). Individuals with high trait anxiety tend to experience more frequent or intense state anxiety episodes, which can in turn elevate stress and reinforce trait anxiety over time. Because state anxiety fluctuates more readily in response to environmental triggers, it may be a more sensitive marker for dietary effects. Indeed, dietary and gut microbiome associations with anxiety appear population-dependent, being more pronounced in subclinical or clinical groups, and diet is more reliably associated with symptom severity (a fluctuating, state-like measure) rather than formal diagnoses (10). If we could 1) determine which dietary and gut microbiome variables are linked to anxiety, and 2) establish a causal link, we could design targeted interventions to help individuals control transient increases in state anxiety and, in the long term, reduce trait anxiety.

Building upon this prior work, the present cross-sectional study investigated how distinct hierarchical dietary levels – including short-term (ST) and long-term intake (LT), nutrient composition, and overall dietary patterns – and gut microbiome features (α, and β-diversity, species and gut-brain modules – GBMs) relate to state anxiety in a cohort of 18–24-year-old females with moderate-to-high trait anxiety. Based on the observed distribution of habitual diet quality, participants were stratified into low- and high-quality clusters using food frequency questionnaire (FFQ)-derived Healthy Eating Index-2020 (HEI-2020) scores, which reflect adherence to the Dietary Guidelines for Americans. The study objectives were to: i) compare state anxiety levels between HEI-2020 clusters (primary hypothesis); ii) to characterize LT and ST dietary intake within each cluster and assess its independent association with state anxiety; iii) examine association of gut microbiome α, and β-diversity with anxiety, iv) identify gut microbiome species and GBMs linearly and non-linearly associated with anxiety; v) assess interactions between anxiety-associated species and GBMs. This study demonstrates that habitual diet quality is significantly associated with state anxiety, and that it may modulate its relationship with short-term dietary intake. Additionally, our findings identify both linear and non-linear associations between anxiety, gut microbiome species, and GBMs. These results support previous evidence linking diet, the gut microbiome, and anxiety, further highlighting several promising microbial targets for microbiome-based interventions aimed at mitigating anxiety.

## 2. Results

### 2.1 Sample characteristics

#### 2.1.1 Metadata by HEI cluster

Fifty-five female participants were enrolled, with nine withdrawals, yielding a final sample of 46. Participants were grouped in two diet clusters (see Methods and **Supplementary materials S1**) with HEI 1 representing lower habitual diet quality (median HEI-2020: 70.9, IQR: 5.1), and HEI 2 higher habitual diet quality (median HEI-2020: 86.3, IQR: 5.1). Both groups underwent the same testing protocol (**Fig. 1**). Briefly, for the primary hypothesis and exploratory analysis, a p-value threshold of 0.05 was adopted for designating statistical significance. All other results are classified as significant with q-values < 0.05, nominal with q between 0.05 and 0.1, and a non-significant trend with q between 0.1 and 0.2 (see Methods). All β coefficient were standardized by scaling (z-scores) predictors and outcome prior to analysis. There were no significant differences in age, BMI, Wechsler Abbreviated Scale of Intelligence (WASI)-II, trait subscale of the State-Trait Anxiety Inventory (STAI-trait), Gastrointestinal Symptom Rating Scale (GSRS), short form of the International Physical Activity Questionnaire (IPAQ-SF), stress subscale of the Depression Anxiety and Stress Scale (DASS)-21, usage of contraceptives, smoking/vaping and self-reported habitual activity (see Methods) between HEI clusters. Full sample description by HEI cluster can be found in **Table 1**.

**Figure 1.**
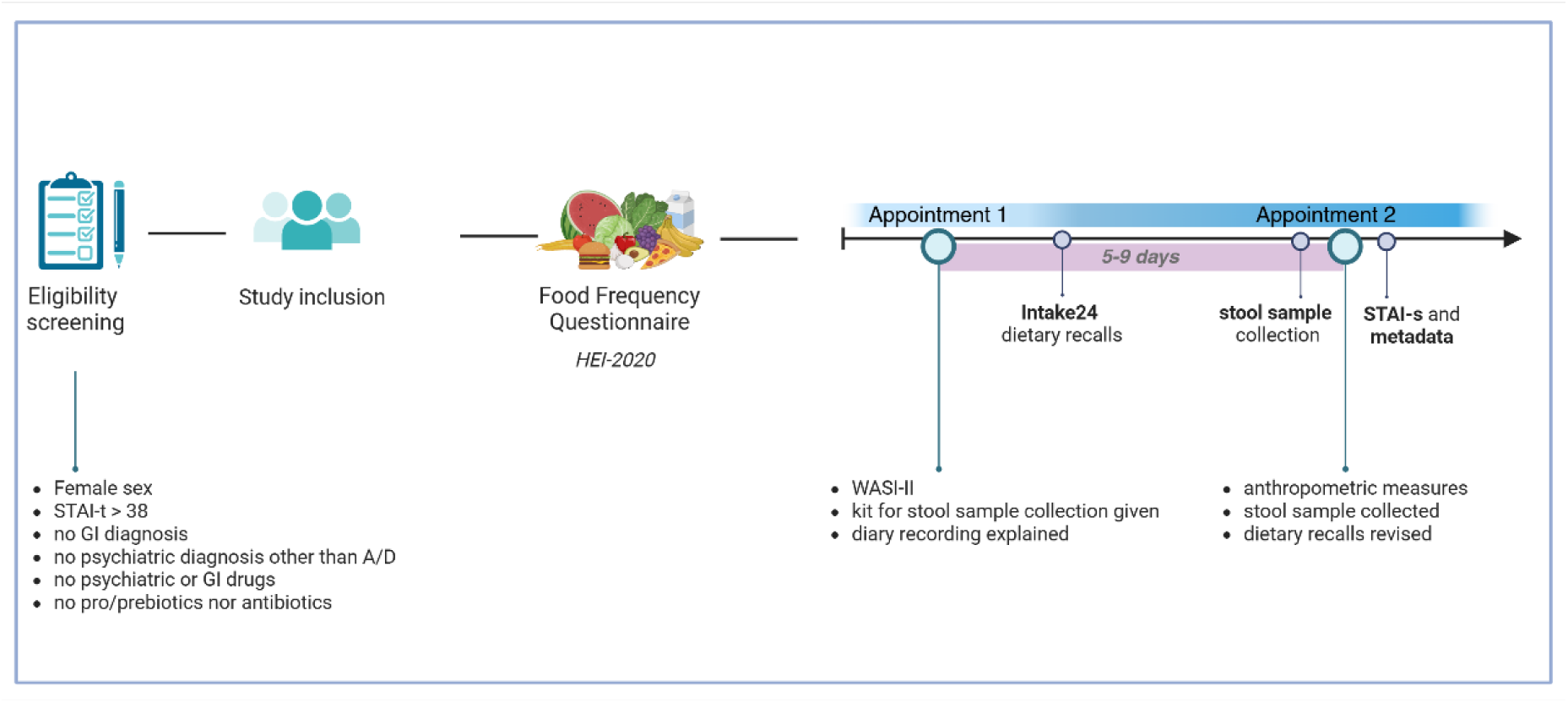
Testing protocol. Following screening, eligible participants completed the Food Frequency Questionnaire (FFQ) to collect long-term dietary data and the HEI-2020 index. Subsequently they i) attended appointment 1 for Intelligence assessment and kit delivery; ii) recorded short-term diet with Intake24; iii) provided a collected stool sample; iv) attended appointment 2 for sample return and anthropometrics; v) completed anxiety and metadata questionnaires. Created with BioRender.com.

**Table 1.**
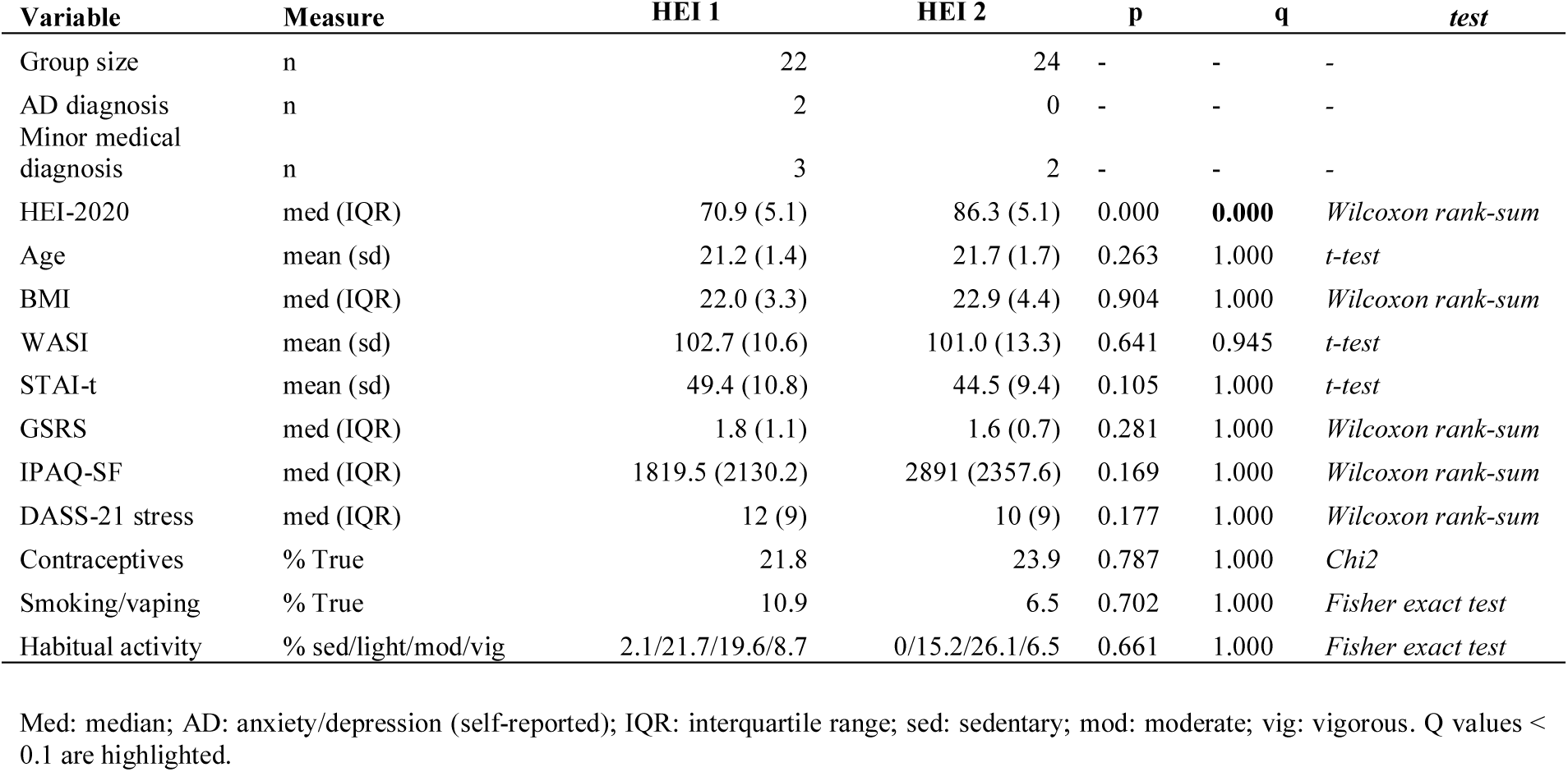
Description of the included sample by HEI cluster.

#### 2.1.2 Diet and gut microbiome phyla by HEI cluster

Higher dietary quality (HEI 2) was driven by higher LT-intake of total and whole fruits, whole grains, fibres, increased unsaturated-to-saturated fat ratio and decreased intake of refined grains and saturated fats (q < 0.05) (**Tables 2** and **3**). Short-term diet of HEI 2 (**Table 4 and 5**) was characterised by i) higher magnesium (q < 0.05), manganese and copper (q < 0.1) intake; ii) significant increase of micronutrients intake driven by higher intake of water-soluble vitamins (increase in vitamin C, thiamine, and folate, q < 0.05); iii) nominal increase of fibre intake and decrease of saturated fats intake (q < 0.1); iv) significant increased intake of fruits (q < 0.05), and a trend in higher intake of red/yellow/green vegetables and beans (q < 0.2); vi) significant increased adherence to a dietary pattern (DP) – defined as Dim. 1 – dominated by fibres, beans and nuts, vegetables and polysaturated fatty acids (PUFAs), along with reduced intake of processed food, saturated fats, meat, cheese and free sugars (**Supplementary materials S2**).

**Table 2.** Summary of HEI subcomponent by HEI cluster. Subcomponents are FFQ-derived and presented as median (interquartile range).

| HEI component | HEI 1 | HEI 2 | p | q |
| --- | --- | --- | --- | --- |
| Vegetables | 5 (0) | 5 (0) | 0.181 | 0.247 |
| **Whole fruits** | 3.94 (1.97) | 5 (0) | 0.004 | **0.014** |
| **Whole grains** | 4.07 (3.77) | 9.07 (2.18) | 0 | **0.002** |
| Green beans | 5 (0) | 5 (0) | 0.345 | 0.407 |
| **Total fruits** | 2.97 (3.3) | 5 (0.29) | 0.006 | **0.015** |
| Dairy | 2.58 (3.63) | 2.62 (6.23) | 0.912 | 0.912 |
| Protein food | 5 (0) | 5 (0) | 0.43 | 0.466 |
| **Protein from seafood \| plants** | 5 (2.28) | 5 (0) | 0.034 | **0.063** |
| **(PUFAs+MUFAs)/SFAs** | 4.46 (2.69) | 9.5 (2.94) | 0 | **0.002** |
| Sodium* | 7.89 (3) | 9.22 (1.95) | 0.094 | 0.153 |
| **Refined grains\*** | 9.32 (2.29) | 10 (0) | 0.002 | **0.01** |
| **Saturated fats\*** | 7.06 (5.13) | 9.18 (2.17) | 0.007 | **0.015** |
| Added sugars* | 10 (0.94) | 10 (0) | 0.19 | 0.247 |

\* a higher score indicates lower intake
PUFAs: polyunsaturated fatty acids, MUFAs: monounsaturated fatty acids, SFAs: saturated fatty acids.
Q values < 0.1 are highlighted.

**Table 3.**
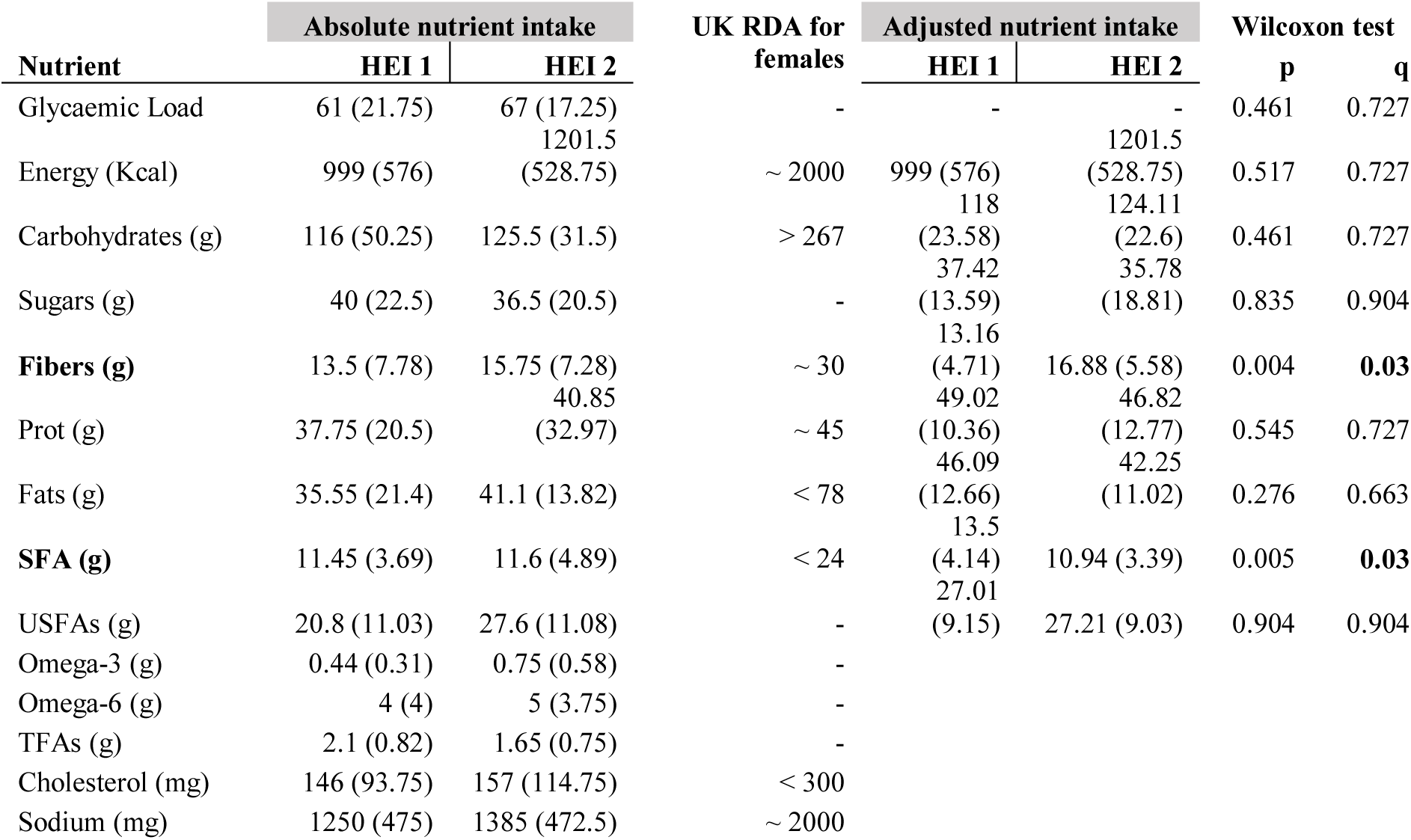
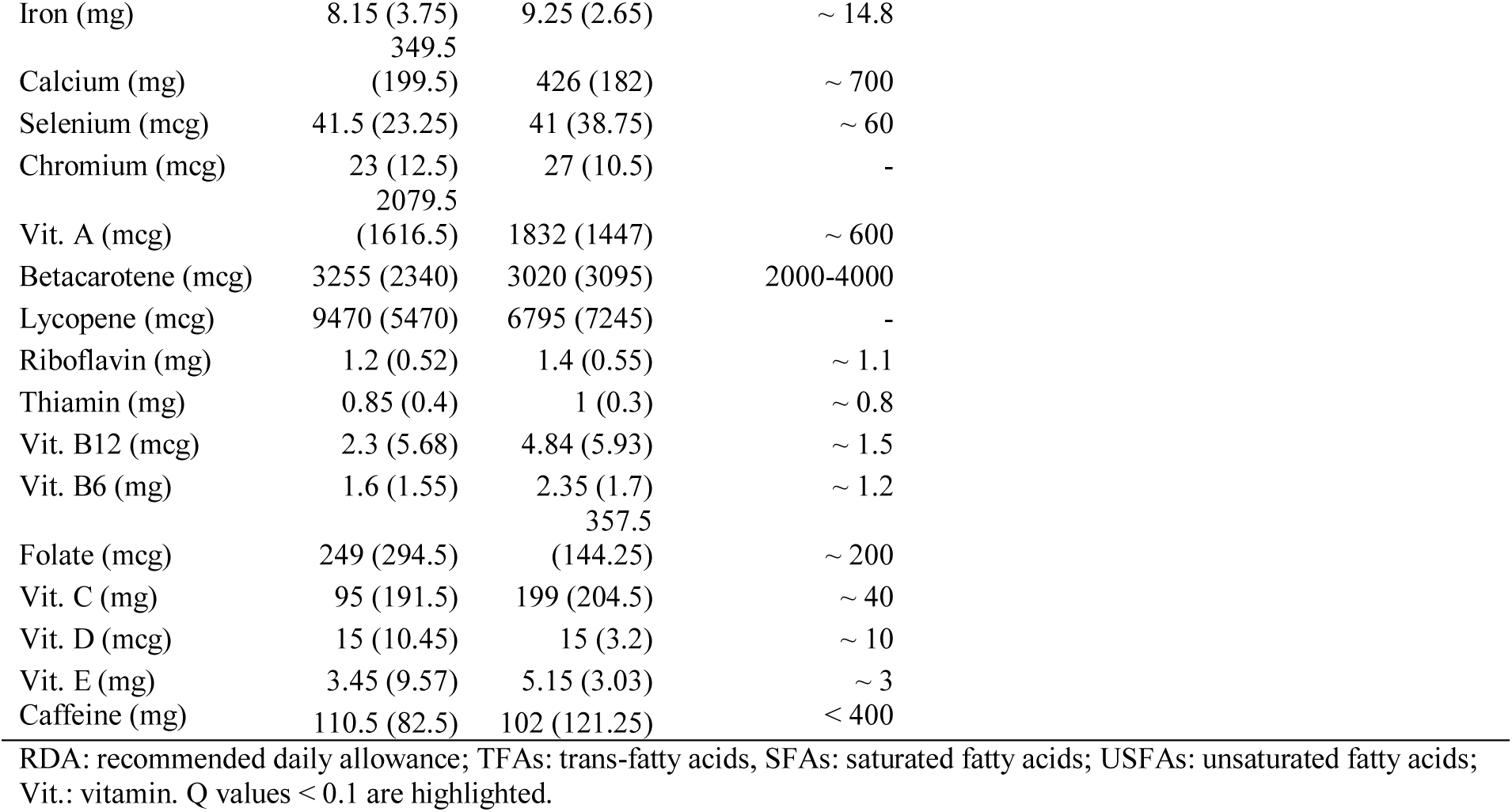
Summary of long-term nutrient intake by HEI cluster. Nutrients are FFQ-derived and presented as median (interquartile range). On the left, all derived raw nutrients are visually compared to the UK Recommended Dietary Allowance (RDA). On the right, only adjusted nutrients used for analysis are presented. Nutrients were adjusted following the residual method.

**Table 4.** Summary of nutrients, nutrient groups, processing level and dietary patterns dimensions by HEI cluster. Data is intake24-derived and presented as median (interquartile range). Dimension (Dim.) 1 and 2 are derived from a Multiple Factor Analysis with details found in supplementary S2.

|  | HEI 1 | HEI 2 | p | q |
| --- | --- | --- | --- | --- |
| **Nutrient** |  |  |  |  |
| Starch (g) | 114.8 (25) | 111.2 (21.6) | 0.783 | 0.854 |
| Non milk extrinsic sugars (g) | 35.9 (25.1) | 37.4 (35.2) | 0.886 | 0.906 |
| **Intrinsic and milk sugars (g)** | 25.7 (17.8) | 39.8 (18.6) | 0.008 | **0.054** |
| **Glucose (g)** | 10.3 (7.8) | 14.9 (6.1) | 0.032 | **0.094** |
| **Fructose (g)** | 9.5 (8.5) | 17.6 (7.6) | 0.008 | **0.054** |
| Maltose (g) | 3.8 (1.1) | 3.1 (1.5) | 0.132 | 0.248 |
| Lactose (g) | 7.2 (8) | 6.5 (6.9) | 0.7 | 0.854 |
| Sucrose (g) | 28.5 (17.5) | 31.3 (24.2) | 0.817 | 0.854 |
| MUFAs (g) | 22.8 (7.1) | 22.4 (4.2) | 0.733 | 0.854 |
| Omega-3 (g) | 1.7 (0.8) | 1.9 (0.8) | 0.097 | 0.198 |
| Omega-6 (g) | 8.6 (3) | 9.3 (3) | 0.397 | 0.642 |
| TFAs (g) | 0.9 (0.4) | 0.7 (0.3) | 0.07 | 0.156 |
| Cholesterol (mg) | 189.7 (134) | 174.3 (152.7) | 0.448 | 0.679 |
| Vitamin A (mg) | 0.5 (0.2) | 0.6 (0.5) | 0.066 | 0.156 |
| Retinol (mg) | 0.2 (0.1) | 0.2 (0.1) | 0.939 | 0.939 |
| **Total carotene (mg)** | 1.4 (1.1) | 2.1 (1.2) | 0.026 | **0.086** |
| Alpha carotene (mg) | 0.1 (0.2) | 0.2 (0.3) | 0.222 | 0.402 |
| **Beta carotene (mg)** | 1.2 (0.9) | 1.9 (1.2) | 0.032 | **0.094** |
| Beta cryptoxanthin (mg) | 0.1 (0.2) | 0.1 (0.2) | 0.7 | 0.854 |
| **Thiamin (mg)** | 1 (0.6) | 1.5 (1.1) | 0.002 | **0.025** |
| Riboflavin (mg) | 1.2 (0.6) | 1.5 (1.1) | 0.106 | 0.208 |
| Niacin (mg) | 16.5 (7.8) | 18.7 (8.6) | 0.361 | 0.629 |
| Tryptophan (mg) | 12.2 (3.2) | 11.9 (4.2) | 0.397 | 0.642 |
| Niacin equivalent (mg) | 31 (10.7) | 30.3 (10.4) | 0.684 | 0.854 |
| **Pantothenic acid (mg)** | 4.7 (2.1) | 5.7 (4.1) | 0.016 | **0.075** |
| **Vitamin B6 (mg)** | 1.3 (0.5) | 1.9 (1.5) | 0.018 | **0.077** |
| **Biotin (mg)** | 0 (0) | 0 (0) | 0.023 | **0.083** |
| **Folate (mg)** | 0.2 (0.1) | 0.3 (0.1) | 0.001 | **0.019** |
| Vitamin B12 (mg) | 0 (0) | 0 (0) | 0.517 | 0.759 |
| **Vitamin C (mg)** | 64.2 (51.5) | 137.6 (165.2) | 0.001 | **0.019** |
| Vitamin D (mg) | 0 (0) | 0 (0) | 0.652 | 0.854 |
| Vitamin E (mg) | 9.6 (3) | 10.1 (6.6) | 0.41 | 0.642 |
| Sodium (mg) | 1604.4 (833) | 1510.2 (458.6) | 0.621 | 0.854 |
| Potassium (mg) | 2059.4 (520.5) | 2245.7 (556.7) | 0.047 | 0.129 |
| Calcium (mg) | 724.3 (249.8) | 671.3 (242.9) | 0.783 | 0.854 |
| **Magnesium (mg)** | 214.7 (58.5) | 255.1 (40.8) | 0.004 | **0.042** |
| Phosphorus (mg) | 1006.8 (230.4) | 1048.6 (214.1) | 0.8 | 0.854 |
| **Copper (mg)** | 0.9 (0.2) | 1 (0.4) | 0.015 | **0.075** |
| Zinc (mg) | 7.1 (1.5) | 7.5 (7.1) | 0.097 | 0.198 |
| Chloride (mg) | 2668.1 (1087) | 2487.7 (953.8) | 0.75 | 0.854 |
| **Manganese (mg)** | 2.3 (1.1) | 3.1 (1.7) | 0.005 | **0.051** |
| Selenium (mg) | 0 (0) | 0 (0) | 0.8 | 0.854 |
| Iodine (mg) | 0.1 (0.1) | 0.1 (0.1) | 0.7 | 0.854 |
| Iron (mg) | 8.2 (3.8) | 10.6 (7.9) | 0.07 | 0.156 |
| Haem iron (mg) | 0.4 (0.3) | 0.3 (0.2) | 0.023 | 0.083 |
| Non haem iron (mg) | 7.6 (3.5) | 10.3 (8.1) | 0.052 | 0.135 |
| **Nutrient group** |  |  |  |  |
| Protein (g) | 61.3 (13.2) | 60.6 (18.3) | 0.7 | 0.77 |
| Fat (g) | 60.4 (15.9) | 59.2 (13.5) | 0.35 | 0.525 |
| Carbohydrate (g) | 178.9 (39.3) | 187.9 (30.9) | 0.397 | 0.556 |
| **Englyst fibre (g)** | 11.3 (3.5) | 14.2 (5.5) | 0.008 | **0.056** |
| **AOAC (g)** | 15.2 (5.5) | 17.8 (6.5) | 0.027 | **0.095** |
| Total sugars (g) | 64.7 (35.7) | 78.4 (28.3) | 0.163 | 0.31 |
| Total FS (g) | 39.2 (25.2) | 40 (30) | 1 | 1 |
| **SFA (g)** | 21.6 (7) | 17.1 (5.1) | 0.013 | **0.075** |
| PUFAs (g) | 10.6 (3.8) | 11.3 (4.1) | 0.35 | 0.525 |
| USFAs (g) | 33.5 (8.7) | 35.1 (8.8) | 0.684 | 0.77 |
| **Micronutrients tot (mg)** | 109.4 (54.8) | 203.1 (167.3) | 0.001 | **0.007** |
| Vitamin fat soluble (mg) | 10.1 (2.9) | 10.9 (6.9) | 0.307 | 0.525 |
| **Vitamin water soluble (mg)** | 100.1 (57.8) | 187.2 (170.7) | 0.001 | **0.007** |
| Minerals tot (mg) | 8505.8 (1641.4) | 8596.5 (2042.7) | 0.7 | 0.77 |
| Minerals salt (mg) | 4306.6 (2033.4) | 4058.8 (1574.9) | 0.733 | 0.77 |
| Minerals no salt (mg) | 4027.2 (1042.6) | 4179.6 (893.8) | 0.15 | 0.31 |
| Omega-6 to omega-3 | 5.6 (1.5) | 5.1 (2.9) | 0.448 | 0.588 |
| SFAs to USFAs | 0.6 (0.3) | 0.5 (0.2) | 0.07 | 0.183 |
| Na to K | 0.7 (0.4) | 0.7 (0.2) | 0.093 | 0.216 |
| **Processing level** |  |  |  |  |
| **Minimal (Kcal)** | 183.2 (160.5) | 278.1 (174.7) | 0.002 | **0.005** |
| **Processed/UPFs (Kcal)** | 821.7 (160.5) | 721.9 (157.4) | 0.005 | **0.005** |

| MFA dimension |  |  |  |  |
| --- | --- | --- | --- | --- |
| Dim 1 | -0.7 (1.4) | 0.2 (1.6) | 0.001 | **0.003** |
| Dim 2 | 0.1 (1.2) | 0 (1.6) | 0.397 | 0.397 |

FS: free sugars; TFAs: trans-fatty acids, SFAs: saturated fatty acids; USFAs: unsaturated fatty acids; Na: sodium; K: potassium; Dim.1 presents high load for fibers, beans, nuts and vegetables; Dim. 2 for lean and non-processed meat. Q values < 0.1 are highlighted.

Consistent with prior human gut microbiota work, Bacillota and Bacteroidota were the dominant phyla across samples (**Fig. 2A**). Prevalence of species and GBMs by HEI cluster can be found in **Supplementary tables ST1**.

**Figure 2.**
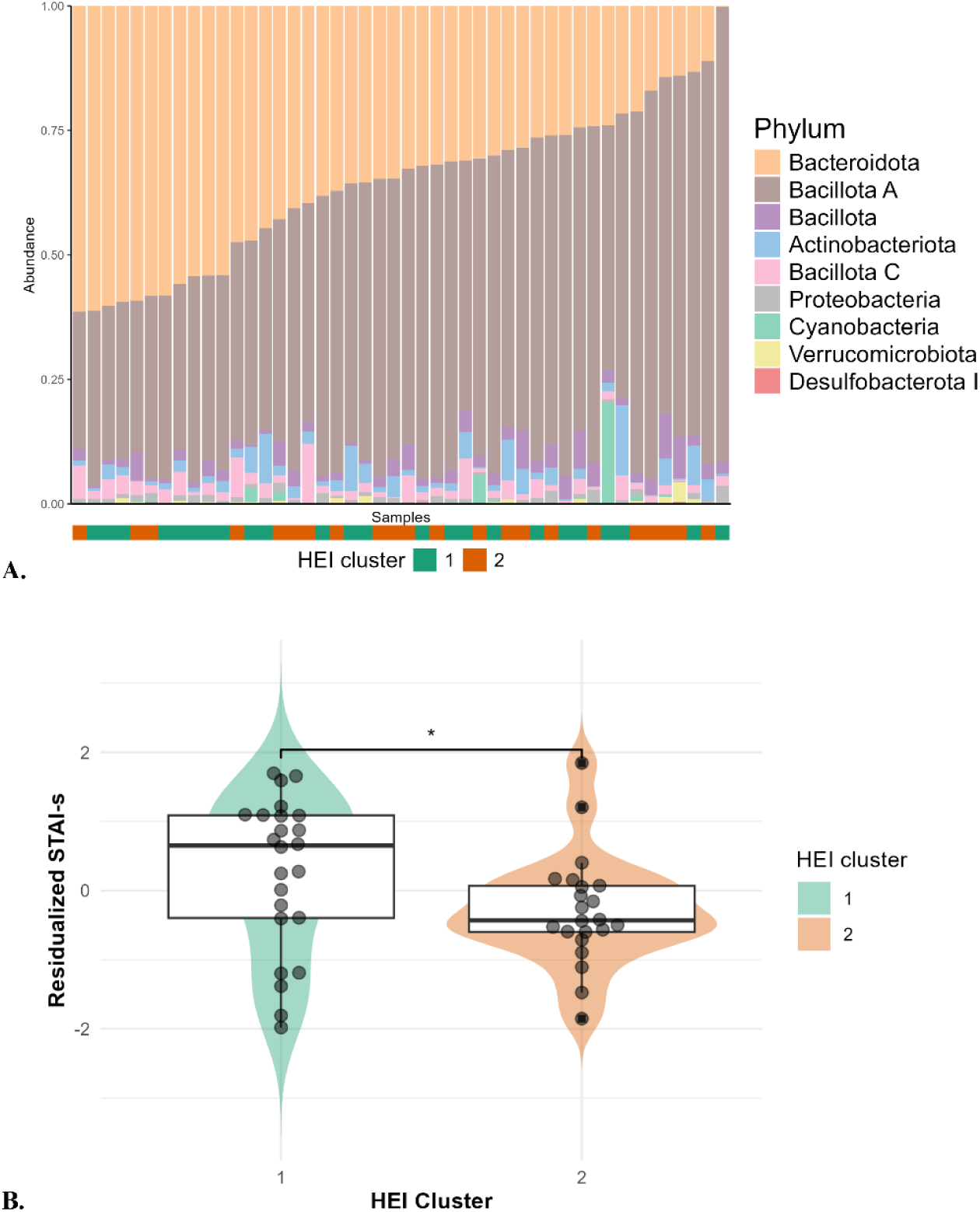
A) Bar chart of the relative phylum-level gut microbiome abundance across participants. Relative phylum-level gut microbiome abundance across participants. The bar chart shows phylum-level abundances within the cohort, ordered by decreasing Bacteroidota. HEI cluster colour codes are displayed below each plot. **B) Violin – Box plots of GSRS-residualized STAI-s score by HEI cluster.** Values of state anxiety (STAI-s) are plotted by HEI cluster. * p value < 0.05. Layout assembled with BioRender.com.

### 2.2 Diet-anxiety associations

#### 2.2.1 Metadata associations with anxiety

First, we assessed whether any metadata were significantly associated with anxiety and found a significant association for stress (DASS-21) and gastrointestinal symptom rating scale (GSRS) (**Table 6**). The anxiety-stress association was expected, as stress is known to act as a trigger and maintenance factor for anxiety. Gastrointestinal symptoms are also associated with stress and anxiety (18) and was controlled for. Accordingly, all subsequent analyses used residualized STAI-s values adjusted for GSRS.

**Table 5.** Summary of food items and food groups by HEI cluster. Data is intake24-derived and presented as percentage of no, low, medium, and high consumption stratified by HEI cluster.

|  | HEI 1 |  |  |  | HEI 2 |  |  |  | p | q |
| --- | --- | --- | --- | --- | --- | --- | --- | --- | --- | --- |
|  | Non-Eaters | Low | Medium | High | Non-Eaters | Low | Medium | High |  |  |
| **Food item** |  |  |  |  |  |  |  |  |  |  |
| **Fruit (%)** | 25 | 58.3 | 12.5 | 4.2 | 4.6 | 31.8 | 59.1 | 4.6 | 0.003 | **0.039** |
| Fruit juice (%) | 25 | 62.5 | 8.3 | 4.2 | 4.6 | 63.6 | 13.6 | 18.2 | 0.025 | 0.162 |
| Tomatoes (%) | 4.2 | 29.2 | 37.5 | 29.2 | 4.6 | 22.7 | 45.5 | 27.3 | 0.879 | 0.879 |
| Tomato puree (%) | 12.5 | 29.2 | 50 | 8.3 | 9.1 | 59.1 | 27.3 | 4.6 | 0.236 | 0.377 |
| Brassicaceae (%) | 37.5 | 29.2 | 33.3 | 0 | 18.2 | 40.9 | 27.3 | 13.6 | 0.131 | 0.243 |
| Yellow/Red/Greens (%) | 0 | 70.8 | 29.2 | 0 | 4.6 | 36.4 | 45.5 | 13.6 | 0.042 | 0.172 |
| Beans (%) | 45.8 | 33.3 | 12.5 | 8.3 | 9.1 | 59.1 | 18.2 | 13.6 | 0.053 | 0.172 |
| Nuts (%) | 41.7 | 29.2 | 25 | 4.2 | 22.7 | 27.3 | 40.9 | 9.1 | 0.11 | 0.238 |
| Other Vegetables (%) | 0 | 29.2 | 58.3 | 12.5 | 0 | 22.7 | 54.5 | 22.7 | 0.388 | 0.504 |
| Beef (%) | 37.5 | 20.8 | 33.3 | 8.3 | 59.1 | 4.6 | 22.7 | 13.6 | 0.503 | 0.545 |
| Poultry (%) | 25 | 29.2 | 33.3 | 12.5 | 54.5 | 0 | 27.3 | 18.2 | 0.464 | 0.545 |
| Cheddar Cheese (%) | 12.5 | 37.5 | 29.2 | 20.8 | 27.3 | 45.5 | 18.2 | 9.1 | 0.082 | 0.213 |
| Other Cheese (%) | 25 | 29.2 | 41.7 | 4.2 | 31.8 | 50 | 9.1 | 9.1 | 0.261 | 0.377 |
| **Food group** |  |  |  |  |  |  |  |  |  |  |
| Processed meat (%) | 37.5 | 29.2 | 20.8 | 12.5 | 54.5 | 31.8 | 9.1 | 4.6 | 0.118 | 0.393 |
| Red meat (%) | 37.5 | 41.7 | 12.5 | 8.3 | 50 | 31.8 | 13.6 | 4.6 | 0.472 | 0.472 |
| All non-processed meat (%) | 12.5 | 25 | 33.3 | 29.2 | 31.8 | 13.6 | 31.8 | 22.7 | 0.295 | 0.393 |
| Fish (%) | 29.2 | 20.8 | 29.2 | 20.8 | 50 | 9.1 | 27.3 | 13.6 | 0.272 | 0.393 |

Q values < 0.1 are highlighted.

**Table 6.**
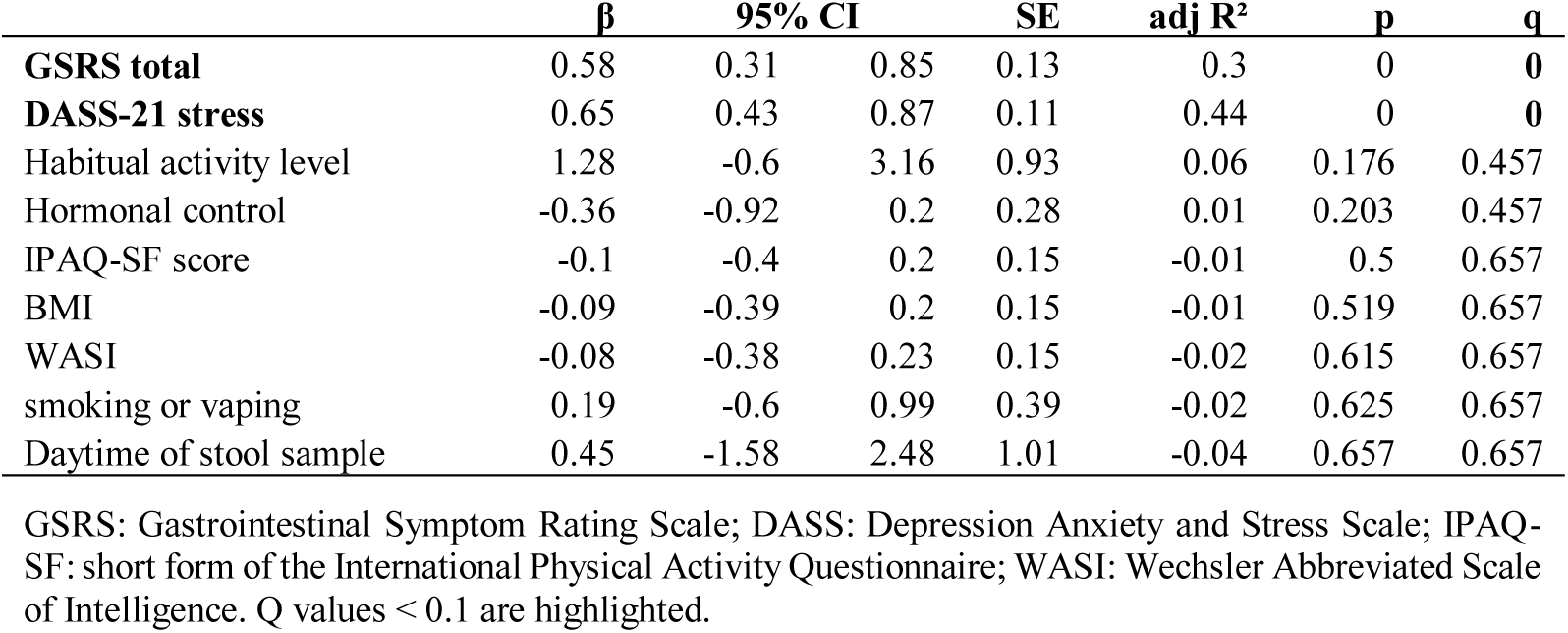
Results from robust regression models on STAI-s and metadata. All beta coefficients are standardized. Habitual activity level, hormonal control, smoking/vaping and daytime of stool sample are categorical predictors.

| | $\beta$ | 95% CI | | SE | adj R<sup>2</sup> | p | q |
| --- | --- | --- | --- | --- | --- | --- | --- |
| **GSRS total** | 0.58 | 0.31 | 0.85 | 0.13 | 0.3 | 0 | **0** |
| **DASS-21 stress** | 0.65 | 0.43 | 0.87 | 0.11 | 0.44 | 0 | **0** |
| Habitual activity level | 1.28 | -0.6 | 3.16 | 0.93 | 0.06 | 0.176 | 0.457 |
| Hormonal control | -0.36 | -0.92 | 0.2 | 0.28 | 0.01 | 0.203 | 0.457 |
| IPAQ-SF score | -0.1 | -0.4 | 0.2 | 0.15 | -0.01 | 0.5 | 0.657 |
| BMI | -0.09 | -0.39 | 0.2 | 0.15 | -0.01 | 0.519 | 0.657 |
| WASI | -0.08 | -0.38 | 0.23 | 0.15 | -0.02 | 0.615 | 0.657 |
| smoking or vaping | 0.19 | -0.6 | 0.99 | 0.39 | -0.02 | 0.625 | 0.657 |
| Daytime of stool sample | 0.45 | -1.58 | 2.48 | 1.01 | -0.04 | 0.657 | 0.657 |

GSRS: Gastrointestinal Symptom Rating Scale; DASS: Depression Anxiety and Stress Scale; IPAQ-SF: short form of the International Physical Activity Questionnaire; WASI: Wechsler Abbreviated Scale of Intelligence. Q values < 0.1 are highlighted.

#### 2.2.2 HEI cluster, short-term and long-term diet associations with anxiety

Robust regressions on HEI clusters revealed that belonging to the HEI 2 (higher diet quality) cluster, compared with HEI 1 was linked to lower anxiety (β = −0.68, SE = 0.29, p = 0.026) (**Fig. 2B**) accounting for approximately 9% of its variance. A nominal negative association was observed between STAI-s and LT fat intake (β = −0.38, SE = 0.15, q = 0.096), with negative trends for both unsaturated and saturated fats (q < 0.2). No associations emerged for ST dietary intake (**Table 7**). Taken together, these findings suggest that habitual dietary quality is the strongest predictor of state anxiety symptoms, with a possible additional contribution of LT fat intake.

**Table 7.**
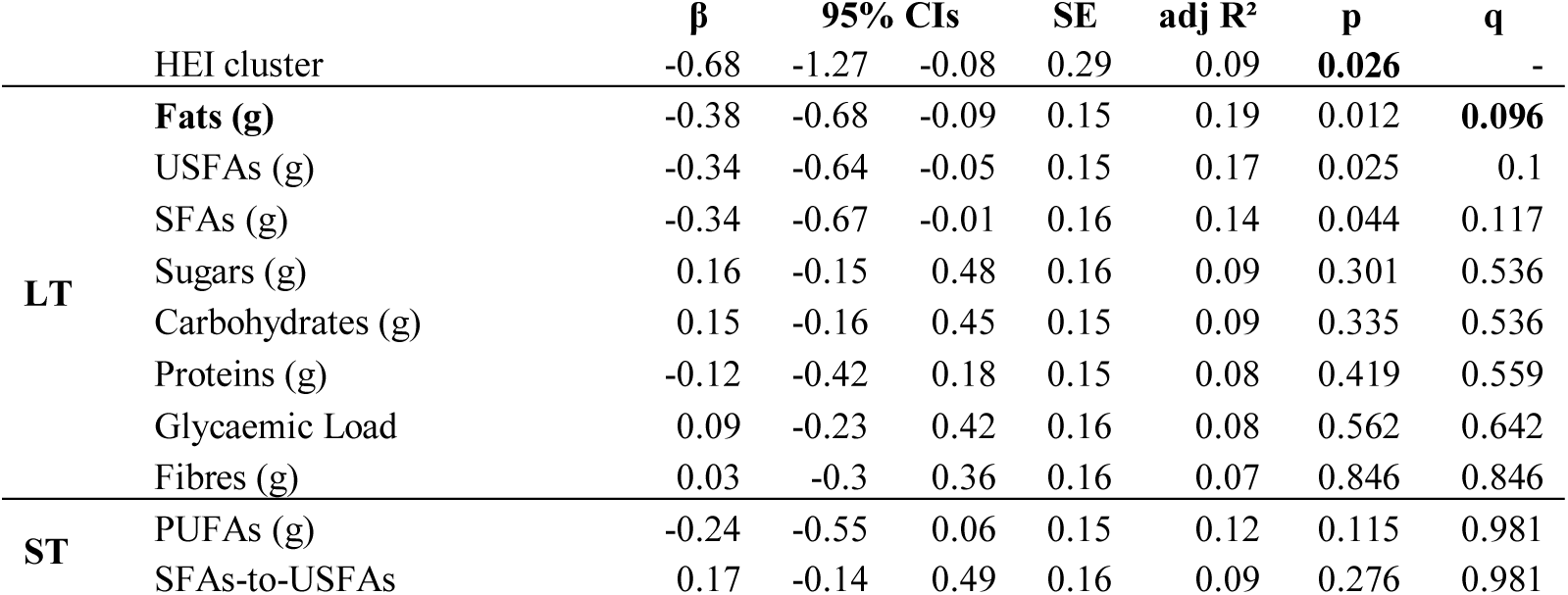
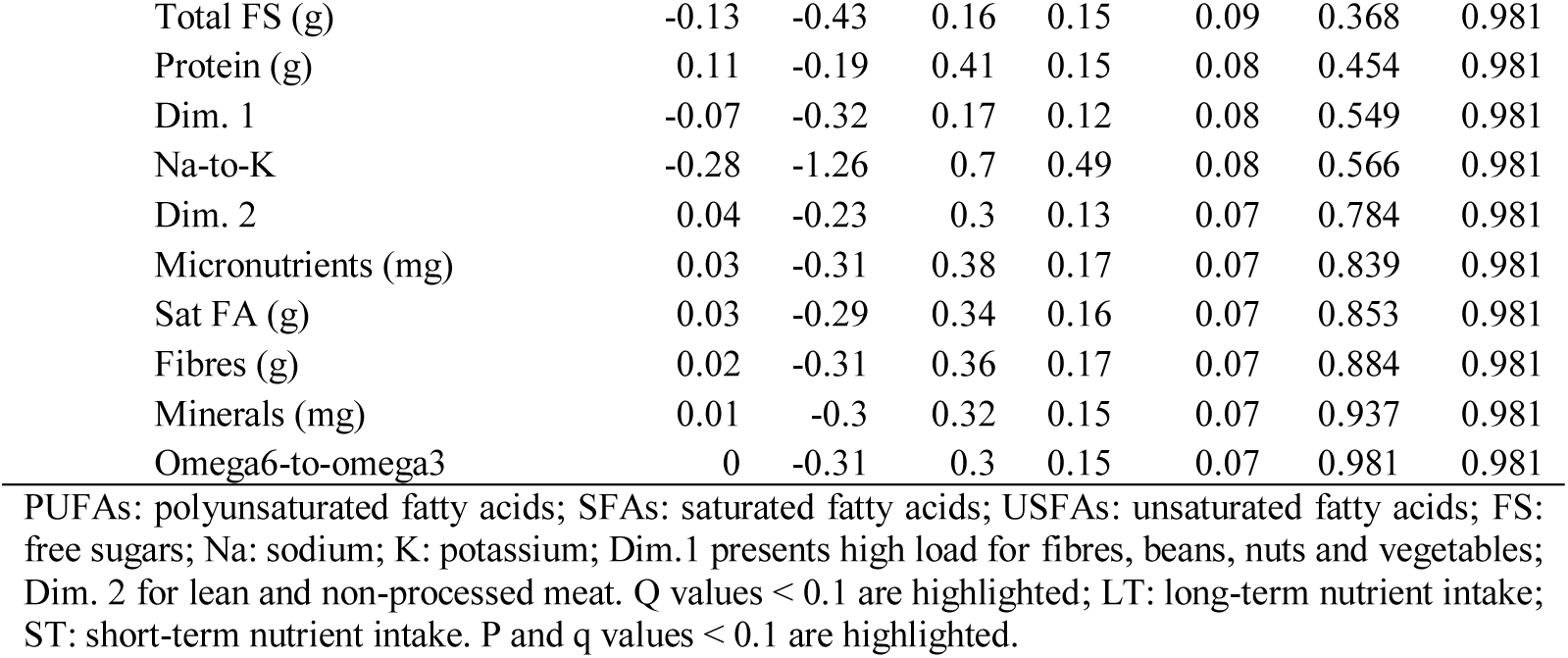
Results from robust regression models on STAI-s and dietary variables. All beta coefficients are standardized. All analysis are performed on residualized STAI-s.

#### 2.2.3 Exploratory analysis of interaction effects between long term and short-term diet and their relationship with anxiety

We did not expect to find negative associations between saturated fat intake and anxiety, nor did we expect the absence of significant effects in the ST dietary intake models. To follow up on these results, we explored whether the association between anxiety and individual dietary variables differed by HEI cluster (i.e., moderation analysis). The rationale was that responses to specific nutrient may depend on the overall dietary context, including long-lasting habits, food choices, processing levels, and nutrient synergy – differences broadly captured by the HEI clusters (see Results 2.1.2). Given the exploratory nature of the analysis, we adopted a raw p value threshold of 0.05 (i.e., not controlling for the false discovery rate). Robust models including STAI-s score as the outcome variable did not show any significant results. We thus repeated the analysis using STAI-s derived categories based on pre-defined cutoff scores (no-to-low anxiety: STAI-s < 38; medium-to-high anxiety: STAI-s ≥ 38) controlling for GSRS. Results from models on HEI cluster and STAI-s categories showed comparable results to continuous models (β = −1.66, SE = 0.68, p = 0.017), indicating that individuals in HEI 2 had 80.3% lower odds of being classified as moderately to highly anxious. In models without interactions, neither LT nor ST dietary intake variables were significant. When including interactions with HEI cluster, we observed a significant main effect of ST PUFA intake (β = - 1.30, SE = 0.65, p = 0.044), and significant interactions for the saturated to unsaturated fatty acids ratio (β_interaction = −3.57, SE = 1.68, p = 0.034) and Dim.1 (β_interaction = 1.66, SE = 0.83, p = 0.045) (**Fig. 3**). The PUFA main effect indicates ∼50% lower odds of being classified as moderate-to-high anxiety per 1-SD higher ST PUFA intake, independent of HEI cluster. For the interaction terms, our results indicate that associations differed by HEI cluster: the saturated to unsaturated ratio was linked to lower odds in HEI 2 but higher odds in HEI 1, higher adherence to Dim.1 related to lower odds in HEI 1 but higher odds in HEI 2.

**Figure 3.**
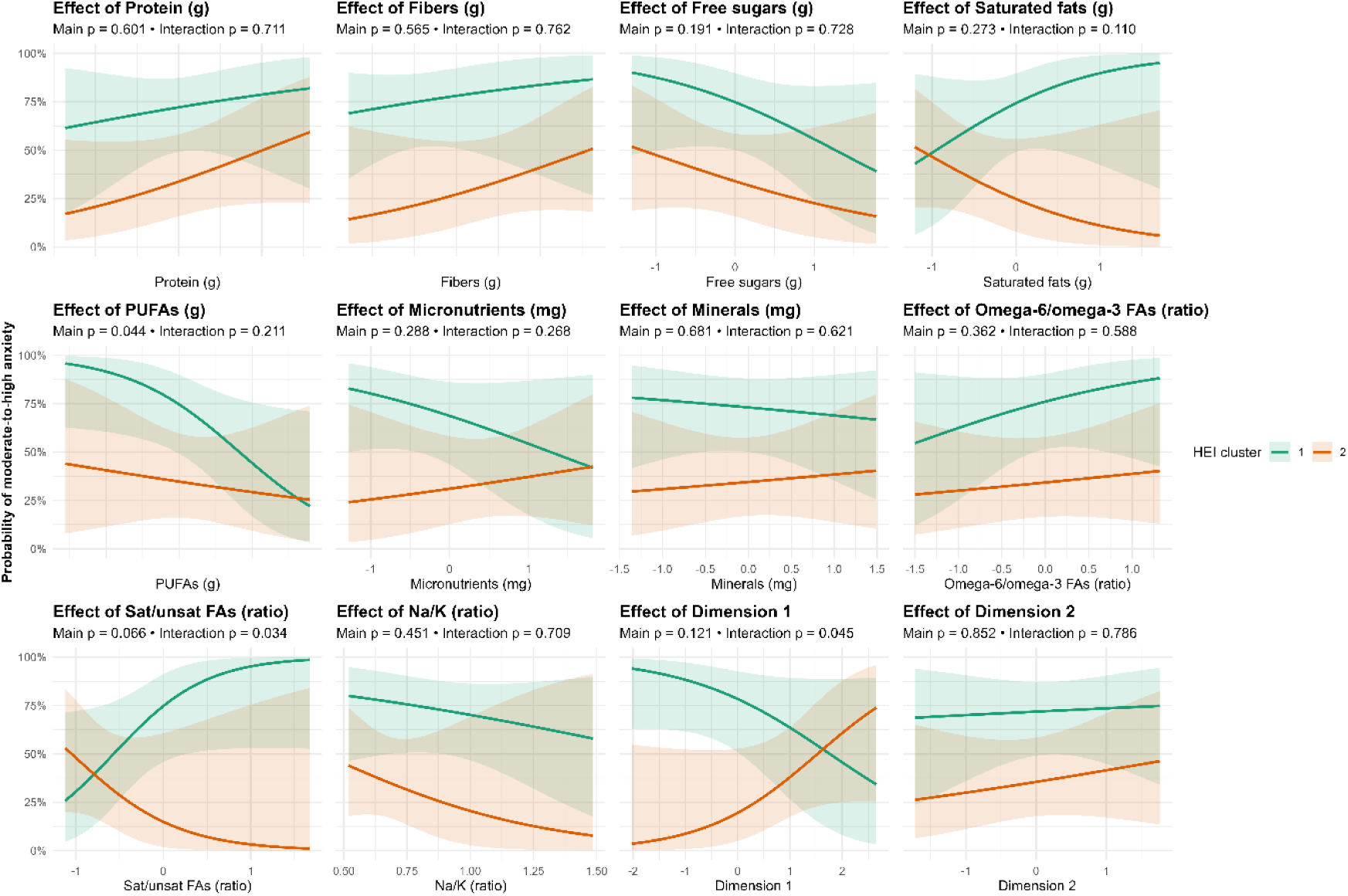
Adjusted predicted probability of medium-to-high anxiety across variation in short-term dietary variables, shown separately by HEI cluster. Curves with 95% confidence intervals, and *p* values are derived from robust logistic regressions controlling for gastrointestinal symptoms (GSRS). All predictors were standardized (z-scored) prior to analysis. FAs: fatty acids.

### 2.3 Linear effects of gut microbiome features

To investigate linear associations between gut microbiome features and anxiety, we first fitted models including only microbiome predictors. We then added habitual diet quality (HEI cluster) to assess whether associations remained after accounting for diet.

#### Results from binary regression models

Robust regression models including absence/presence of commensal gut species indicated a significant anxiety increase associated to *Flavonifractor plautii* presence (q = 0.000), a trend toward higher anxiety in the presence of *Ruminococcus B gnavus*, and decreased anxiety when *Faecalibacterium prausnitzii J* and *E* were present (q = 0.157-0.196). Standardized β coefficients ranged from –1.15 to 1.2, with 95% CIs excluding zero, and the models explained 16–25% of the variance in anxiety (adjusted R² = 0.16–0.25) (**Table 8**, **Fig. 4**). After including HEI cluster as covariate, β coefficients attenuated by ∼ 0.2 and q values increased (q > 0.3). Finally, belonging to HEI 2 cluster was associated with a trend toward lower probability of *F. plautii* presence (std β = −1.68, SE = 0.88, q = 0.136), and higher probability of *F. prausnitzii E* (std β = 1.47, SE = 0.41, q = 0.136). Overall, these results suggest diet-confounded effects where the presence of *F. prausnitzii J* and *E* is linked to lower transitory anxiety symptoms, whereas the presence of *F. plautii* and *R. B gnavus* to higher anxiety symptoms.

**Figure 4.**
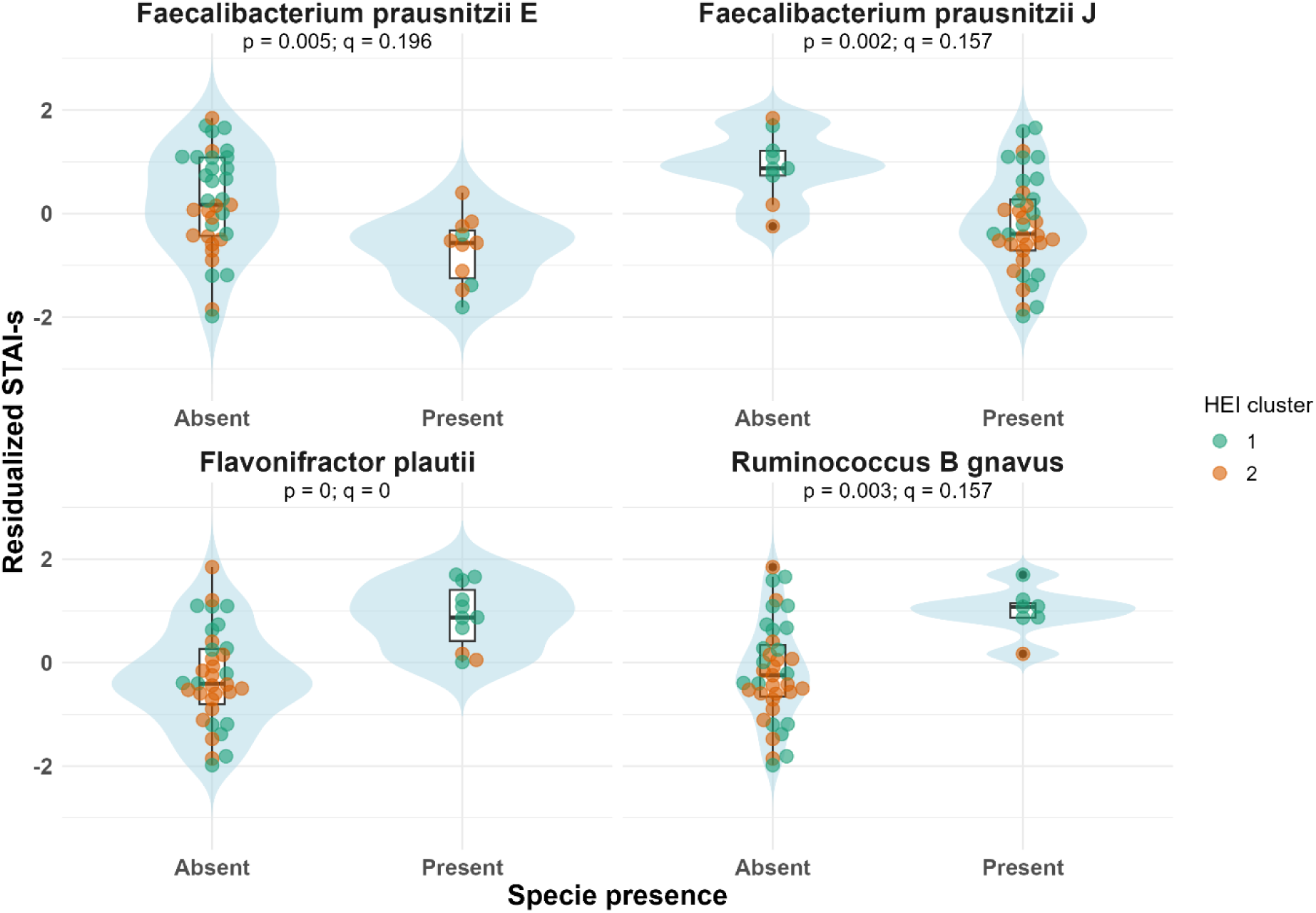
Associations between absence/presence of species and anxiety. Only species with q values < 0.2 are shown. Violin – Box plots display GSRS-residualized STAI-s scores by species presence, with individual points coloured by HEI cluster.

**Table 8.** Results from robust regression models on STAI-s and binary gut microbiome species. The reference category was absence. All beta coefficients are standardized. All analysis are performed on residualisd STAI-s.

| Species | $\beta$ | 95% CIs | | SE | adj R<sup>2</sup> | p | q |
| --- | --- | --- | --- | --- | --- | --- | --- |
| *Flavonifractor plautii* | 1.2 | 0.57 | 1.83 | 0.31 | 0.25 | 0 | **0** |
| *Faecalibacterium prausnitzii* *J* | -1.15 | -1.87 | -0.44 | 0.35 | 0.19 | 0.002 | 0.157 |
| *Ruminococcus B gnavus* | 1.2 | 0.42 | 1.98 | 0.39 | 0.17 | 0.003 | 0.157 |
| *Faecalibacterium prausnitzii* *E* | -0.98 | -1.65 | -0.32 | 0.33 | 0.16 | 0.005 | 0.196 |

Q values < 0.1 are highlighted.

#### Results from continuous regression models

We fit robust models for commensal species and GBMs clr-transformed abundances among subjects where they were detected (i.e., samples with at least one read count for that feature) to assess whether increasing abundances were linked to STAI-s. Zeros before clr transformation were handled by the Bayesian-multiplicative method (see Methods). We observed positive, nominally significant associations for an unclassified *Bacteroides* species (unclassified 31, std β = 0.49, q = 0.082) and *Bilophila wadsworthia* (std β = 0.67, q = 0.082), and a positive trend for *Bacteroides thetaiotaomicron* (std β = 0.50, q = 0.164). The respective models accounted for 21, 39, and 22% of the outcome variance (**Table 9**, **Fig. 5**). In the HEI cluster-adjusted models, the association for *B. thetaiotaomicron* was no longer nominally significant, while the associations for the unclassified 31 specie and *B. wadsworthia* were became trend-level (q = 0.164). In the *B. wadsworthia* model, the explained variance remained 39%, and the HEI cluster term itself became non-significant. These results highlight a positive link between these species and anxiety symptoms and suggest that the HEI cluster and species abundance may capture overlapping portions of the explained anxiety variance.

**Figure 5.**
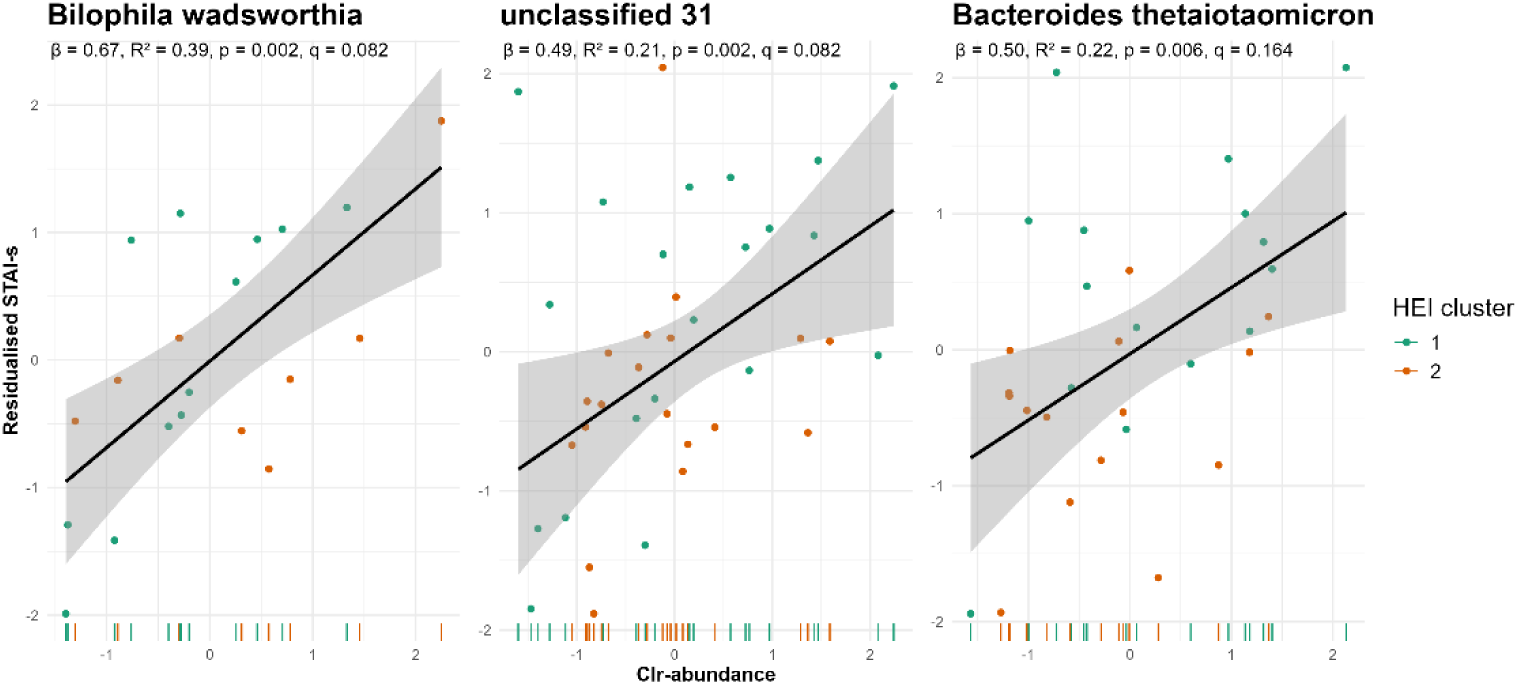
Associations between species abundance and anxiety. Only species that showed q values < 0.2 are shown. Scatterplots of residualized STAI-s (y-axis) against clr-abundance (x-axis) per each species, with individual points coloured by HEI cluster.

**Table 9.** Results from robust regression models on STAI-s and gut microbiome species and gut-brain modules abundance. All beta coefficients are standardized. All analysis are performed on residualisd STAI-s.

| | | $\beta$ | 95% CIs | | SE | adj R<sup>2</sup> | p | q | N |
| --- | --- | --- | --- | --- | --- | --- | --- | --- | --- |
| Species |  |  |  |  |  |  |  |  |  |
|  | unclassified 31 | 0.49 | 0.19 | 0.78 | 0.15 | 0.21 | 0.002 | 0.082 | 39 |
|  | Bilophila wadsworthia | 0.67 | 0.28 | 1.07 | 0.19 | 0.39 | 0.002 | 0.082 | 20 |
|  | Bacteroides thetaiotaomicron | 0.5 | 0.15 | 0.84 | 0.17 | 0.22 | 0.006 | 0.164 | 30 |
| GBMs |  |  |  |  |  |  |  |  |  |
| MGB053 | Butyrate synthesis II | -0.32 | -0.57 | -0.08 | 0.12 | 0.13 | 0.01 | 0.187 | 46 |
| MGB021 | GABA synthesis II | -0.51 | -0.89 | -0.13 | 0.19 | 0.14 | 0.011 | 0.187 | 43 |

Results from GBMs models revealed a negative trend between STAI-s and butyrate synthesis II (std β = −0.32, SE = 0.12; q = 0.187) and GABA synthesis II (std β = −0.51, SE = 0.19, q = 0.187) pathways (**Table 9**, **Fig. 6A**). For both associations q values increased (q > 0.3) after accounting for HEI cluster. No associations were found between HEI cluster and anxiety-associated GBMs. Finally, no significant associations were observed between anxiety and α - diversity metrics, nor Bacillota-to-Bacteroidota ratio.

**Figure 6.**
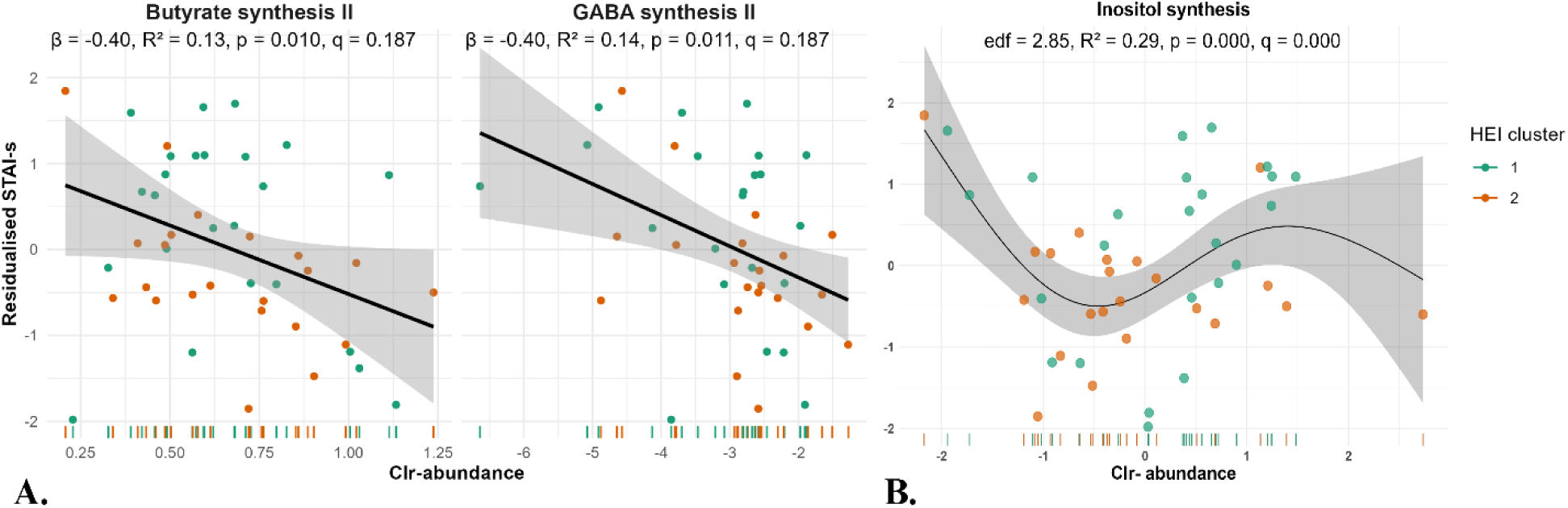
A) Associations between gut-brain modules and anxiety. Only GBMs that showed q values < 0.2 are shown. Scatterplots of residualized STAI-s (y-axis) against clr-abundance (x-axis) per each species, with individual points coloured by HEI cluster. **B) Non-linear association between inositol synthesis and anxiety.** The scatter plot shows individual participants (coloured by HEI cluster) with a smooth fitted curve; the shaded area indicates 95% confidence intervals. Inositol synthesis abundance is expressed as standardized clr-transformed abundance. Layout assembled with BioRender.com.

### 2.4 Non-linear effects of gut-brain modules

To investigate any non-linear effects not captured by the robust regressions, we fitted adjusted Generalised Additive Models (GAMS) for each GBM among subjects with non-zero counts for a given feature. The inositol synthesis module showed a significant U-shaped association (edf = 2.85, q = 0.000) explaining 29% of the variance in anxiety. The spline breakpoint was between −1 and 0 clr-abundance, with STAI-s decreasing as inositol synthesis increases at its lower values and increasing at its higher values (**Fig. 6B**). Although the spline appears to show an S-shape, wide confidence bands on the right tail reflect data sparsity and uncertainty. A sensitivity analysis of the inositol association was performed, removing 2 influential points (hat > 3 × mean hat value), which confirmed a significant U-shape with the same breakpoint pattern. A linear trend was observed between the GABA synthesis II module and anxiety (edf = 1, q = 0.136) with 13% accounted variance. When including HEI cluster in the models, inositol synthesis reached nominal significance (edf = 2.8, q = 0.068) with HEI cluster becoming a trend (q = 0.19) with overall variance explained remaining unchanged. GABA synthesis II remained nominal (edf = 1, q = 0.128) and both butyrate and propionate synthesis II modules showed linear trends (edf = 1, q = 0.128) (**Supplementary Table ST2**). Overall, these data suggest a linear relationship between butyrate, propionate, and GABA synthesis, partially confounded by diet quality, and a non-linear relationship between inositol synthesis and anxiety, independent of diet quality.

### 2.5 Associations between species and gut-brain modules

To visually assess interactions between anxiety-associated species and GBMs, we built a clustered heatmap based on standardized β coefficients from robust models adjusted for HEI cluster (see Methods) (**Fig. 7**). Significant and trend-level associations were observed between *F. prausnitzii E* and butyrate (std β = 1.02; q = 0.016) and propionate synthesis (std β = 0.81; q = 0.116); *Bacteroides* unclassified 31 sp. and inositol synthesis (std β = 0.33; q = 0.117); and *B. wadsworthia* and butyrate (β: −0.56; q: 0.189) and GABA synthesis (β: −0.56; q: 0.063). Most Bacillota species, except *F. plautii,* showed positive associations with butyrate synthesis. *F. plautii*, *B. wadsworthia* and the *unclassified 31* sp. were negatively associated with GABA synthesis. Butyrate and GABAsynthesis (and, to a lesser extent, propionate synthesis) clustered together across coefficients, whereas inositol synthesis covaried more independently.

**Figure 7.**
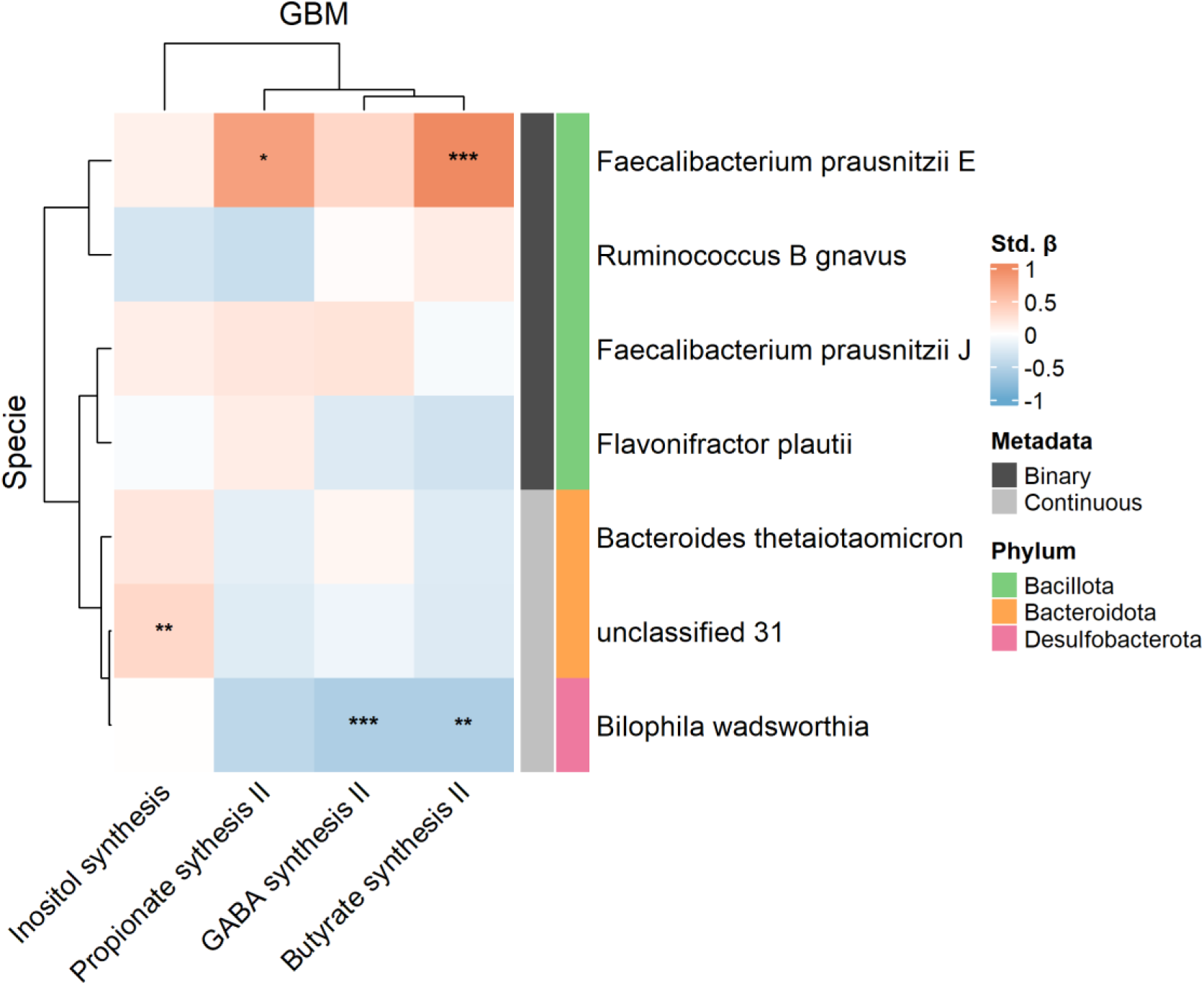
Clustered heatmap of associations between anxiety-associated species and gut-brain modules (GBMs). The heatmap displays standardized β coefficients from robust linear models, depicting the relationship between each anxiety-associated gut microbial species (rows) and GBMs (columns). Cells colors indicate effect magnitude (std. β), and asterisks denote significance levels: *** *q* < 0.05, ** 0.05 ≤ *q* < 0.1, * 0.1 ≤ *q* < 0.2. Rows are annotated by model type (binary or continuous regressions) and phylum-level taxonomy. Rows and columns are hierarchically clustered based on coefficient similarity.

### 2.7 β-diversity not associated with anxiety

To test whether individuals with similar overall gut microbiome profiles have similar anxiety scores, we applied a Microbiome Regression-Based Kernel Association Test (MiRKAT) on species and GBMs Aitchison distance, adjusting for HEI cluster. Results showed a trend in significance for species with little variance explained (R^2^: 0.02, q: 0. 125) and no evidence of association for GBMs (R^2^: 0.002, q: 0.41) (**Supplementary Figures S1**, **S2**), suggesting that that β-diversity is not a strong predictor for anxiety levels on a continuous scale, in a subclinical sample.

## 3. Discussion

### Habitual diet was associated with anxiety

In our cross-sectional study, habitual dietary quality (HEI-2020) was the strongest predictor of state anxiety, outperforming both ST nutrient intake and nutrient based measures. We found that higher habitual diet quality was linked to *F. prausnitzii* presence and *F. plautii* absence. No significant variation in gut microbiome functional profiles was observed across HEI-2020 clusters. The few and weak associations may reflect small effect sizes and the limited sample size. Also, because HEI-2020 assigns fixed weights to components with unequal microbial relevance (e.g., fibre-rich components vs seafood-based components), it may not fully capture dietary effects on the gut microbiota. Future studies could address this by developing a microbiome-informed dietary index, with weights informed by each component’s impact on microbial metabolism, to more accurately characterize long-term diet–microbiome relationships.

There were no significant associations with ST dietary intake variables after false discovery rate correction, but we were interested in exploring potential interaction effects between habitual and ST diet in relation to anxiety. We performed a moderation analysis to investigate these potential interactions. A negative association was observed between anxiety and PUFAs, which have been repeatedly linked to better mood and anxiety outcomes (19,20). Among individuals with lower habitual diet quality, we found the expected patterns: higher saturated-to-unsaturated fat ratios were associated with greater anxiety, whereas a fibre-rich and plant-based food pattern was associated with lower anxiety. Strikingly, the opposite trend emerged among individuals with higher habitual diet quality, suggesting context-dependent effects of nutrient intake and anxiety. One possible explanation relates to dietary composition and food matrix effects. Participants in the HEI 2 cluster consumed less saturated fat, more fibre and plant-based foods, and an overall more nutrient-dense, minimally processed diet. In such a context, fat intake is more likely to be derived from whole-foods and may interact differently within a less processed food matrix. Another explanation involves psychological and behavioural factors. Occasional consumption of indulgent foods may support mental health and flexibility among individuals with generally healthy diets. Individuals with high trait anxiety and high habitual diet quality may display heightened dietary consciousness, sometimes adopting rigid “healthy eating” behaviours consistent with orthorexia (i.e., pathological preoccupation with healthy eating) (21). Notably, even non-pathological or “healthy” orthorexia has been positively associated with stress and anxiety. Taken together, these exploratory findings highlight the need to consider both nutritional context and psychological dynamics when interpreting food–anxiety relationships. Without accounting for habitual dietary patterns and food-related behaviours, it may be difficult to clearly capture the signal linking diet to anxiety.

The lack of significance between ST dietary intake and anxiety may be explained by the study’s limited sample size. Our study was powered for small-to-medium effects: smaller effects, especially amid relatively high levels of inter-individual heterogeneity, might have been missed (9). However, larger cohorts may increase the risk of false positives and spurious associations driven by unknown confounders or collider variables (22). We thus emphasise effect sizes and clinical relevance rather than statistical significance alone. The Minimal Clinically Important Difference (MCID) represent the smallest within-person change perceived as beneficial, but it could also be valuable to adopt a similar metric in cross-sectional studies to contextualize the practical importance of scientific findings. No MCID is established for STAI-s in healthy or clinically anxious populations, hence we prioritised standardised coefficients and recommend developing an STAI-s MCID (e.g., prior surgical cohorts suggest ≈8 points) (23).

Overall, our data suggest that habitual diet is the primary dietary determinant of anxiety variability. We show for the first time that short-term diet associations with anxiety may be context-dependent and strongly encourage future studies to account for the multifactorial nature of eating behaviour. This approach should improve biomarker discovery and account for a substantial fraction of inter-individual variability in nutritional psychiatry.

### Several gut microbiome features were associated with anxiety

Taxa associated to anxiety in our study broadly align with prior research. Most – but not all – studies report an inverse association between *Faecalibacterium* and anxiety, with analyses typically conducted at the genus level (10,24,25). *Facealibacterium* is a major butyrate producer (consistently with results from our species-GBMs association models), which is known to reduce inflammation and support gut-barrier integrity (26,27). In murine models of chronic low-grade inflammation*, F. prausnitzii* reduces colonic cytokines and serotonin levels (28) – all mechanisms implicated in anxiety pathophysiology (29).

Consistent with our findings, Jiang et al. observed enrichment *of R. B gnavus* in patients with generalized anxiety, even in the absence of overt symptoms (30). By contrast, Ganci et al. reported the inverse association in a female cohort (31). Such discrepancies may reflect host-specific factors and strain- and context- dependent immunomodulation: for example, the same strain (i.e., ATCC 29149) can be either pro- or anti-inflammatory depending on which surface polymer it expresses (32). Individual diet could also modulate *R. B gnavus* effect on health: low polysaccharides intake may increase mucin degradation and compromise gut barrier function. A similar process could be at play with *B. thetaiotaomicron* – a polysaccharide degrader able to break down the mucosal layer (33). *B. thetaiotaomicron* can also promote the growth of the sulfidogenic pathobiont *B*. *wadsworthia* by deconjugating bile acids and releasing taurine (33), and both bacteria were associated with increased anxiety in our study. *B*. *wadsworthia* produces hydrogen sulphide (H_2_S), which may have protective effects at physiological levels but, in excess, can disrupt the gut epithelium, promote oxidative stress, and contribute to systemic and neuroinflammation (34). H₂S may also influence the gut–brain axis directly, potentially affecting vagal signalling and neurotransmitter release (34). Prior research has suggested a role for H_2_S in the pathogenesis of neurodegenerative disorders, such as Alzheimer’s disease (34), but further research is needed to define how these mechanisms may contribute to anxiety pathophysiology.

The presence of *F. plautii* was positively associated with anxiety symptoms, which we have not seen reported elsewhere. Earlier work linked *F. plautii* enrichment to affective disorders and post-traumatic neuropsychiatric sequelae, sometimes alongside increased *R. B gnavus* (35–37). *F. plautii* is a flavonoid degrader that exerts anti-inflammatory effects in metabolic syndrome and colitis mouse models (38,39), yet its role in psychiatric outcomes is poorly understood. Culture-based work has isolated a strain from a related species (93.4% similar to *F. plautii*) that requires the presence of GABA for growth and subsequent studies suggested *F. plautiii* could be a GABA-consuming bacterium (40–42). Depressive, posttraumatic, and anxiety disorders share dysregulated stress-response systems and altered GABAergic signalling, with lower GABA levels frequently reported. While luminal GABA is unlikely to cross the blood-brain barrier and rich the central nervous system, potential *F. plautii*-induced reduction in luminal GABA could still influence host GABAergic signalling by affecting vagal afferents or epithelial-enteric pathways and contribute to symptom expression (43). Trials supplementing with oral GABA report stress-reducing effects, though autonomic contributions (sympathetic versus parasympathetic) are mixed, and higher doses are usually required to detect central-nervous-system changes (43).

We tested for non-linear associations between gut microbiome features and anxiety and observed a U-shape relationship for the inositol synthesis pathway. Given that many physiological processes operate within an optimal range and that several health outcomes are sensitive to microbiome balances (44,45), complementing linear with non-linear modelling may help to identify important relationships that might otherwise remain undiscovered. Inositol is a crucial molecule involved in insulin signalling, oxidative defence, cell membrane integrity, and neurotransmission (46). In a in randomized controlled trial, inositol supplementation at ≥ 12g/day was shown to reduce symptoms in patients affected by panic disorders, with comparable efficacy to fluvoxamine (46,47). However, we speculate that elevated microbial inositol-synthesis potential may result in trade-offs that coincide with lower SCFAs **(Fig. 9**), although this idea requires further exploration and validation. Other host and dietary factors may also influence these non-linear dynamics. For example, excessive luminal glucose and simpler sugars may hinder inositol absorption (46,48), resulting in elevated intestinal concentrations but reduced host availability. Furthermore, inositol can serve as a source of arachidonic acid (46), which, under certain conditions (e.g., in the presence of environmental stressors), may contribute to inflammatory responses. Inositol signalling may also indirectly affect GABAergic circuits via its modulation of serotoninergic, noradrenergic, and dopaminergic receptors (46). A preclinical epileptic model showed an increase in expression of GABA-A receptor subunits α4 and γ2 following myo-inositol supplementation. Overexpression of the α4 subunit following long- and short-term exposure to positive GABA-A modulators (e.g., progesterone-derived neuroactive metabolite - allopregnanolone) has been associated with anxiogenic effects (49,50). Although little is known about the interaction between inositol and GABA signalling in humans, the mechanisms discussed above may partially explain the non-linear association we observed, which warrants further investigation.

Taken together, our data and the available literature suggest the presence of *R. B gnavus* and abundance of *B*. *wadsworthia* are key player in the gut microbiome- anxiety associations and that *F. prausnitzii* might have protective effects possibly through butyrate production. Our data also suggests *F. plautii* and microbial GABA synthesis potential as key players in the gut-brain axis, making them plausible biomarkers and mechanistic targets that requires further investigation. Importantly, inositol might act as an underlying player by regulating neurotransmitters signalling within an optimal range to exert beneficial effects. While the pathways remain speculative, testing these mechanisms of action and clarify whether any effect primarily happen at the gut- or brain-level would advance our understanding of anxiety pathophysiology. Importantly, defining the interplays with the stress-immune axis would yield a holistic mechanistic understanding that could guide future research and intervention development.

### Limitations

Our study had a few notable limitations. First, the modest sample size limited covariate adjustments and precluded a fully specified biopsychosocial model (51). We therefore used a GSRS-residualized approach and prioritized diet as key covariate for gut microbiome models. Second, the sample size did not allow us to fit a multivariate model including all gut microbiome features simultaneously. Hence, their combined contribution will need to be further explored in larger, independent cohorts. Third, the cross-sectional design precludes causal inference yet provides mechanistic insights and identifies candidate microbial biomarkers for further investigation.

## Conclusions

To the best of our knowledge, this is the first study focusing on young females with moderate to high-trait anxiety to jointly analyse short- and long-dietary intake, taxonomic and functional gut microbiome features, and state anxiety. We identified habitual diet as the major dietary driver of anxiety variability and hypothesise that long-term diet reshapes microbial community composition and functioning, thereby conditioning short-term exposure-response relationships. We found that *R. B gnavus*, *F. prausnitzii*, *F. plautii*, and *B*. *wadsworthia* explained some of the observed variation in state anxiety. Our findings further suggest associations between state anxiety and SCFAs pathways (propionate and butyrate) and GABA metabolism, and a more nuanced, non-linear relationship between microbial inositol production and anxiety. Disentangling these dynamics will require longitudinal, multilevel modelling approaches that account for inter-individual heterogeneity. We recommend adopting a personalized, multilevel, and systems-based framework to guide participant-tailored trials and accelerate translation of microbiome-mediated interventions in nutritional psychiatry.

## 4. Methods

### 4.1 Participants and study design

Participants were recruited via flyers, social media, and mail-outs at the University of Surrey (UK). Screening took place online via the Qualtrics platform (https://www.qualtrics.com/). We included females aged 18–24 years with medium-to-high trait anxiety, as measured by the State-Trait Anxiety Inventory-trait (STAI-t) with scores ≥ 38 (17). Exclusion criteria were: (i) self-reported psychiatric diagnosis other than anxiety or depression; (ii) use of psychiatric medications; (iii) self-reported gastrointestinal diagnosis (e.g., gastritis, IBS); and (iv) use of antibiotics or probiotics or fructo- and galacto-oligosaccharides supplements within the previous three months. After reading the study information, participant gave consent by ticking an online checkbox consent form and received a monetary reward after study conclusion. All participants provided informed consent prior to taking part in the study. We calculated that a minimum sample size of 39 and 48 was required with one, and two predictors, respectively, to reach a small-to-medium effect size of f^2^= 0.2 (52) with α of 0.05 and 80% power. Ethical approval was granted by the University of Surrey Ethics Committee (Reference number: FHMS-22-23-052).

### 4.2 Testing design

Participants attended two in-person appointments. At the first appointment, they completed the Wechsler Abbreviated Scale of Intelligence-II (WASI-II) and received: verbal and written instructions on how to accurately record i) their dietary intake using Intake24; and ii) a stool self-collection kit. Participants were instructed to collect their sample just before the second appointment and after completing their dietary recording. Appointment 2 took place 5-9 days after appointment 1, based on the participants’ availability. During this appointment, the self-collected stool sample was handed-in, and anthropometrics (height, weight) measures were taken. Participants were then instructed to complete an online Qualtrics survey – for outcome and metadata collection (Section 2.3.2) – within the following day (although some participants took longer than 1 day to complete the entries). Spot checks were conducted on Intake24 dietary entries to ensure data quality.

### 4.3 Data collection

#### 4.3.1 Dietary intake assessment

A web-based 67-item Food Frequency Questionnaire (FFQ) (Quadram Institute Bioscience and SafeCape Software Solutions (53,54)) was used to assess habitual diet quality using the HEI-2020 of the previous 6 months. The HEI-2020 total score and its 13 components were automatically derived with higher scores reflecting greater adherence to the Dietary Guidelines for Americans (55). The FFQ also provided estimated LT daily nutrient intakes.

Short-term dietary intake of nutrients and food items was assessed using the web-based 24-hour recall platform Intake24 (https://intake24.co.uk/). Intake24 uses data from the UK Nutrient databank, including the McCance & Widdowson Composition database. It is based on a multiple-pass recall approach, consisting of a free retrospective recall followed by detailed questions – a system with accuracy comparable to face-to-face interviews (56). The intake24 platform was chosen because i) it was iteratively developed for optimal usability and engagement (57); ii) it has been validated across multiple population, including 11–24-year-olds (58–60); iii) it outperformed the alternative food log app (61). Participants were given a unique, anonymous access link to the platform and instructed to complete recalls on three non-consecutive weekdays when feasible (e.g., Monday, Wednesday, and Friday) and one weekend day, providing a representative measure of their weekly intake. Compliance was monitored in real time, with reminder emails sent when recalls were missed.

#### 4.3.2 Psychological assessment and metadata

An online Qualtrics survey was sent straight after the second appointment, including the i) STAI-state (STAI-s) to assess state anxiety on a continuous scale from 20 to 80 (62); ii) DASS-21 to assess the subcomponent DASSI-21 stress (ranging 0-42) (63); iii) GSRS with total score reported as mean of the original 1-to-7 scale (64–66); iv) the IPAQ-SF, measuring physical activity during the previous 7-days as metabolic-equivalent minutes per week. Participants were also asked to self-report i) usage of contraceptives and of any medications, smoking/ vaping, and habitual physical activity level (sedentary, light, moderate, vigorous).

#### 4.3.3 Stool sample collection, DNA extraction and sequencing

Participants self-collected their stool samples at home using ZymoResearch DNA/RNA Shield Fecal Collection Tube (R1137). Samples were then stored at 4C, DNA extracted using the QIAamp PowerFecal Pro DNA kit and stored at −20C before being shipped in dry ice to the Quadram Institute Bioscience. Genomic DNA was normalised to 5 ng/µl with EB (10mM Tris-HCl). A miniaturised reaction was set up using the Illumina Nextera DNA Flex Library Prep Kit (Illumina Catalogue No 20018704). 0.5 µl Tagmentation Buffer 1 (TB1) was mixed with 0.5 µl Bead-Linked Transposomes (BLT) and 4.0 µl PCR grade water in a master mix and 5 μl added to a chilled 96 well plate. 2 µl of normalised DNA (10 ng total) was pipette mixed with the 5 µl of the tagmentation mix and heated to 55 ⁰C for 15 minutes in a PCR block. A PCR master mix was made up using 10 µl KAPA 2G Fast Hot Start Ready Mix (Merck Catalogue No. KK5601) and 2 µl PCR grade water per sample. 12 µl of this mastermix was added to each well to be used in a 96-well plate. 1 µl of 10µM 10bp Unique Dual Indexes were added to each well. Finally, the 7 µl of Tagmentation mix was added and mixed. The PCR was run with 72⁰C for 3 minutes, 95⁰C for 1 minute, 14 cycles of 95⁰C for 10s, 55⁰C for 20s and 72⁰C for 3 minutes. The libraries were quantified using the Promega QuantiFluor® dsDNA System (Catalogue No. E2670) and run on a GloMax® Discover Microplate Reader. Libraries were pooled following quantification in equal quantities. The final pool was double-SPRI size selected between 0.5 and 0.7X bead volumes using sample purification beads (Illumina® DNA Prep, (M) Tagmentation (96 Samples, IPB), 20060059). The final pool was quantified on a Qubit 3.0 instrument and run on a D5000 ScreenTape (Agilent Catalogue No. 5067-5579) using the Agilent Tapestation 4200 to calculate the final library pool molarity. Samples were sent to Novogene (Novogene (UK) Company Limited, 25 Cambridge Science Park, Milton Road, Cambridge, CB4 0FW, United Kingdom) to be run along with sample names and index combinations used. Faecal samples were shotgun sequenced (paired end) using an Illumina NovaSeq X (25B lanes) at Novogene.

#### 4.3.4 Genome reconstruction, taxonomic and functional profiling of metagenomes

MG-TK (https://github.com/hildebra/mg-tk) was used to process metagenome raw reads, assemble metagenomes, and reconstruct and dereplicate metagenomic-assembled genomes (MAGs) (67). Raw metagenomic reads were quality filtered using sdm v1.63 with default parameters (68). Kraken2 (69) was used to remove human contaminated reads. Host-filtered metagenome reads were assembled using MEGAHIT v1.2.9 (70) with default parameters and reads then back-mapped to the assembly using strobealign (71). Genes were predicted with Prodigal v2.6.1 with parameters “-p meta” (72) and a gene catalogue was de novo created by clustering these at 95% nucleotide identity using MMseqs2 (73). Matrix operations on the gene catalogue were carried out using rtk (74).

From metagenomic assemblies, MAGs were calculated using SemiBin2 (75) and their completeness and contamination estimated using CheckM2 (76). Dereplication into MGS (metagenomic species) was carried out via clusterMAGs (https://github.com/hildebra/clusterMAGs), relying on both SemiBin2 high quality MAGs (>80% completeness, <5% contamination) and canopy clusters (https://github.com/hildebra/canopy2) (77). Function in the gene catalogue were annotated as described in Frioux *et al.* (78) using Diamond blastp mapping against the eggNOG database (79)(80) and KEGG orthologs (81). Brain-related metabolic functional potential was assessed with a previously published database of manually curated gut-brain modules (GBMs, with each corresponding to a single neuroactive compound production or degradation process).

### 4.4 Data preprocessing and feature engineering

#### 4.4.1 Dietary data

Based on the bimodal distribution of HEI-2020 scores, participants were stratified into two clusters representing lower (HEI 1) and higher (HEI 2) diet quality. Clustering was performed using a k-means algorithm with Euclidean distance and assessed with the elbow method and silhouette analysis (see **Supplementary Material S1**). HEI components were used for sample characterization and group-level comparison of habitual diet. FFQ-derived nutrient values were adjusted for total energy intake using the residual method (82).

Single day recalls from Intake24 were averaged per participant to derive ST dietary intake. Multilevel variables of interest included: i) nutrient groups and ratios; ii) processing level of food consumption; iii) overall dietary patterns. Food items and groups were used to compute processing categories and overall patterns, and to describe dietary differences between HEI clusters. Nutrient groups and ratios were created by aggregating individual nutrient values and adjusted using the residual method. Food groups were generated by aggregating individual food items, as classified by the Intake24 platform, and both food items and groups were standardized to grams per 1000 Kcal of energy intake. Food variables with more than 50% of non-consumers were excluded. Given the skewed and sparse distribution typical of food intake data, we applied k-medoids clustering to discretize intake into four ordinal levels: non-consumer, low, medium, and high intake. Non-consumers were excluded from clustering to maintain a conceptual distinction between consumption vs. non-consumption. Clustering was then performed on non-zero intake values (k = 3) using Euclidean distance. Each participant was assigned a medoid-based category value, representing the median intake within their cluster (whereas non-eaters were assigned to 0), which was later used in Multiple Factor Analysis (MFA) to reflect inter-level distances. The processing level of food consumption was computed by categorizing 749 standard sub-food items from Intake24 according to the NOVA classification system (83). Items in NOVA category 4 were classified as UPFs, and intake was then expressed as energy-standardized values (Kcal per 1000 kcal). To identify overall dietary patterns, we applied MFA using nutrient groups, food processing levels, and medoid-encoded food items as distinct blocks. This allowed us to define underlying factors integrating different dietary levels and extract DPs. Full details are provided in **Supplementary materials S2**.

For statistical analysis, the skewness of all independent variables was assessed, and variables with skewness greater than (absolute) 1 were log-transformed. All variables were then scaled. Severe multicollinearity between nutrient groups/ratios, and processing levels was assessed using the variance inflation factor (VIF). When VIF exceeded 10 (84) a correlation matrix was computed to identify redundant variables (r > 0.8) (84). In each highly correlated pair, one variable was removed based on:

1. relevance to the study aim (e.g., Englyst fibre was retained over AOAC fibres as the latter includes lignin - not fermentable by gut microbes and thus not directly contributing to SCFAs production) and
2. comprehensiveness (e.g., total micronutrients was preferred over subclass distinctions). A final set of 12 short-term dietary intake predictors, including 2 MFA-derived dimensions, was retained for downstream analysis.

#### 4.4.2. Gut microbiome data

##### Taxonomy

Taxonomic features of interest were species. Phylum-level data were retained only for descriptive purposes and Bacillota-to-Bacteroidota ratio calculation. Species were rarefied to the smallest read count to compute presence/absence and α-diversity, including Shannon, Inverse Simpson, and observed richness (R package rtk 0.2.6.1) (85). Relative abundances were used for descriptive purposes. For all other continuous analysis, species were filtered based on mean relative abundance ≥ 0.01% (86,87). Two complementary approaches were adopted:

1. Binary modelling: rarefied species with prevalence between 15% and 85% were retained and encoded as binary variables (presence/absence), yielding 157 species used in subsequent models.
2. Continuous analysis: species with prevalence > 20% (133 species) were retained. Following a Compositional Data approach (88,89), zeros were replaced via a Bayesian-multiplicative method - which preserves overall data structure (89,90) - by using the R function cmultRepl (method = “CZM”, output = “p-counts”) from the zCompositions (1.5.0-4) package (91). We then applied centred log-ratio transformation (clr).

Finally, we assessed β-diversity using the Aitchison distance (Euclidean in clr-space) and visualized sample clustering via Principal coordinates Analysis (PCoA). For all gut microbiome-specific processing steps the mia (1.14.0) R package was used (92).

### Gut Brain Modules

KEGG orthologs (KOs) were rarefied to compute α-diversity indices. Gut-Brain Modules were identified from raw KOs using the R package Omixer-RPM (93,94) with coverage threshold of 30% (95) and were then pre-processed as described for species. Given the low sparsity of GBMs data, only compositional analysis was carried out. Finally, β-diversity was assessed and visualized as described above.

### 4.5 Descriptive statistics

#### 4.5.1 Metadata and dietary differences between HEI clusters

For descriptive purposes, data was summarised by HEI clusters. The distribution of continuous variables was assessed using violin and QQ plots, and the Shapiro–Wilk test. Normally distributed metadata were summarised as mean (sd) and compared between groups using independent t-tests. Non-normally distributed metadata were summarised as median (IQR) and compared using the Wilcoxon rank-sum test. Dietary data was also described per each HEI cluster. For consistency and robustness, all adjusted continuous variables (nutrients and dietary processing levels) were summarised as median (IQR) and compared between clusters using the Wilcoxon test, regardless of distribution. FFQ-derived raw average intake across the cohort is also presented alongside recommended daily allowances for females for reference. Given generally lower validity coefficients for micronutrients compared to macronutrients and other nutrients in FFQ validation studies, micronutrients were used for descriptive purposes but not included in statistical analyses, to prioritise robustness and interpretability of results (96). Categorical variables were reported as proportions (%). Categorical metadata was compared between groups using Fisher’s exact test or the chi-squared test, as appropriate. Discretized food item and group categories were treated as ordinal variables and compared using the Cochran-Armitage test to assess graded consumption patterns across HEI clusters.

#### 4.5.2 Gut microbiome composition

Overall sample composition was visualized at phylum level with mia::plotAbundance. Relative prevalence of species and GBMs within each HEI cluster was also calculated and reported as relative proportions.

### 4.6 Statistical Analysis

#### 4.6.1 Metadata and diet-anxiety regression models

Because some of our nutritional and biological variables exhibited mild skewness and outliers even after data cleaning, and given our sample size, we used robust statistical models throughout with setting = KS2014 (97,98). This approach ensured to apply consistent analysis across the entire pipeline, while guarding against assumption violations, outliers, and influential points. Standardized coefficients were calculated to allow interpretation of effect size across variables. Robust linear regression models via the R function lmrob (robustbase 0.99-4-1)(99) were applied. We first examined whether collected metadata variables were associated with STAI-s scores and used residualized STAI-s values (adjusted for GSRS) in all subsequent analyses (see Results section). To assess if HEI clusters predicted different anxiety scores, we modelled STAI-s ∼ HEI cluster. The same models were then applied to LT- and ST-nutrient intakes adjusting for HEI cluster. Results from the full-adjusted models are reported, including standardized regression coefficient (β), standard error (SE), 95% confidence intervals (CIs), p value, and adjusted R². Based on the results from models including LT- and ST dietary variables, we conducted exploratory analysis to test whether their association with anxiety differed between HEI clusters. Robust logistic models (robustbase::glmrob) were applied with anxiety measured as categorical dependent variable (no-to-low anxiety: STAI-S < 38; medium-to-high anxiety: STAI-S ≥ 38).

#### 4.6.3 Gut microbiome-anxiety regression models

To assess the relationship between gut microbiome features and anxiety, we fitted robust regression models with STAI-s as outcome. First, we examined presence/absence of individual species (STAI-s ∼ binary species). Subsequently, HEI was included as a covariate to control for dietary effects. The same pipeline was applied to continuous species and GBMs abundances (restricted to subjects in which the feature was present i.e., count > 0, and to features with N ≥ 20) to test whether increasing abundance of a given feature was associated with STAI-s, to α diversity metrics and Bacillota-to-Bacteroidota ratio. Binary models were not applied to GBMs because prevalence was > 85% for all of them. Finally, we assessed the relationship between anxiety-associated species/GBMs and HEI cluster.

#### 4.6.4 Gut microbiome-anxiety generalized additive models

To assess non-linear relationship between STAI-s and continuous GBMs abundances in subjects where present, we fit penalised regression splines using the R package mgcv (1.9.3) (100). A basis dimension of *k* = 4 was chosen to balance flexibility and prevent overfitting. We adopted the scaled-*t* family to down-weight outliers. We estimated the smoothing (i.e., λ penalty of curvature) and dispersion parameters by restricted maximum likelihood which yields less-biased estimates of variance components than maximum likelihood and is recommended for modest samples (101,102). HEI cluster was subsequently included as linear covariate. Effective degrees of freedom (EDF) – indicating the approximate amount of “bends” in each smooth – adjusted R^2^ and p values were extracted from each model. Pointwise 95% confidence bands were computed using the draw function and graphically displayed. Generalized Additive models were not applied to species due to high data sparsity, hence lack of power.

#### 4.6.5 Data integration

To assess interactions between anxiety-associated species (binary and continuous) and GBMs, we constructed a hierarchical clustering heatmap. Robust regression models were fitted for each GBM with specie as predictor and HEI cluster as covariate. Standardized β coefficients and were extracted, q values computed for significance annotation. The resulting effect size matrix was visualized using the ComplexHeatmap R package (103). Clustering was performed using Pearson correlation (to group features by similar directionality patterns) with complete linkage, which is less sensitive to outliers.

4.6.6 β diversity analysis

We tested the association between overall gut microbial community profiles and STAI-s by using MiRKAT via the R package MiRKAT (1.2.3) (104). MiRKAT fits a kernel-machine regression where the outcome (STAI-s) is modelled as a function of a pairwise microbiome similarity matrix, testing whether more similar gut microbiome profiles tend to have more similar anxiety scores and allowing for covariates adjustment (104). We derived the similarity matrix from the Aitchison dissimilarity matrix via the R function D2K and then applied MiRKAT with permutation. We then reported i) R² (i.e., proportion of outcome similarity explained by microbiome similarity); ii) q-value.

#### 4.6.7 Multiple testing control and software

We adopted a p-value threshold of 0.05 for the primary hypothesis testing and exploratory analysis. To control for multiple testing within each subsequent analysis, we adjusted p-values using the False Discovery Rate with the Benjamini-Hochberg procedure. Given the limited sample size and the large number of tests performed, we adopted a q-value threshold of 0.20 with q < 0.05 here defined as significant, q between 0.05 and 0.1 as nominal, and q between 0.1 and 0.2 as trend. All data processing and analysis were performed in R (version 4.4.3), using R studio (version 2025.05.0). The following packages were further used for data processing and visualization: dplyr (1.1.4), tidyverse (2.0.0), factoextra (1.0.7), cluster (2.1.8), FactoMineR (2.11), e1071 (1.7-16), car (3.1-3), DescTools (0.99.60), rstatix (0.7.2), ggplot2 (3.5.1), ggbeeswarm (0.7.2), ggpubr (0.6.0), miaViz (1.14.0), scater (1.34.0), patchwork (1.3.0).

## 5. Data availability

All sequencing data generated as part of this study are publicly available in the European Nucleotide Archive (ENA) under the study accession PRJEB105368. All code used for data processing and analysis is publicly available in GitHub (https://github.com/m-basso/2025_dma_cross_sectional). Data used for analysis are provided in the **Supplementary Tables ST3-ST8**.

## 6. Additional files

Supplementary Materials S1-S2-FS1-FS2

File format: .docx

Supplementary Tables ST1-ST8

File format: .xlsx

## 7. Authors contribution

MB: Conceptualization, Data curation, Formal analysis, Investigation, Project administration, Methodology, Visualization, Writing – original draft. FH: Data curation, Methodology, Software, Resources, Writing – review & editing CW: Project administration, Data curation. DB: Data curation, Writing – review & editing. RM: Supervision, Writing – review & editing. MBar: Methodology, Supervision, Validation, Writing - Review & Editing SMG: Methodology, Validation, Supervision, Writing -review & editing. KCK: Conceptualization, Supervision, Funding acquisition, Writing – review and editing.

## 9. Declarations

### Ethics approval and consent to participate

Ethical approval for this study was granted by the University of Surrey Ethics Committee (Reference number: FHMS-22-23-052). Before participation, all participants signed an informed consent.

### Consent for publication

Not applicable

## Data Availability

All sequencing data generated as part of this study are publicly available in the European Nucleotide Archive (ENA) under the study accession PRJEB105368. All code used for data processing and analysis is publicly available in GitHub (https://github.com/m-basso/2025_dma_cross_sectional). Data used for analysis are provided in the Supplementary Tables ST3-ST8.

## Acknowledgments

We kindly thank Will Boulton for the upload of the FASTQ files used in this study.

